# A randomized controlled trial of alpha phase-locked auditory stimulation to treat symptoms of sleep onset insomnia

**DOI:** 10.1101/2024.01.10.24301120

**Authors:** Scott Bressler, Ryan Neely, Ryan Yost, David Wang

**Affiliations:** Elemind Technologies, Inc., Cambridge, MA, United States

## Abstract

Sleep onset insomnia is a pervasive problem that contributes significantly to the poor health outcomes associated with insufficient sleep. Auditory stimuli phase-locked to slow-wave sleep oscillations have been shown to augment deep sleep, but it is unknown whether a similar approach can be used to accelerate sleep onset. The present randomized controlled crossover trial enrolled adults with objectively verified sleep onset latencies (SOLs) greater than 30 minutes to test the effect of auditory stimuli delivered at specific phases of participants’ alpha oscillations prior to sleep onset. During the intervention week, participants wore an electroencephalogram (EEG)-enabled headband that delivered acoustic pulses timed to arrive anti-phase with alpha for 30 minutes (Stimulation). During the Sham week, the headband silently recorded EEG. The primary outcome was SOL determined by blinded scoring of EEG records. For the 21 subjects included in the analyses, stimulation had a significant effect on SOL according to a linear mixed effects model (p = 0.0019), and weekly average SOL decreased by 10.5 ± 15.9 minutes (29.3 ± 44.4%). These data suggest that phase-locked acoustic stimulation can be a viable alternative to pharmaceuticals to accelerate sleep onset in individuals with prolonged sleep onset latencies.

## Introduction

Insufficient sleep is a global problem that is associated with an increased risk of premature mortality and a long list of adverse health conditions^1^. Insomnia is the most prevalent sleep disorder among adults, and more than 60% of individuals experiencing insomnia report difficulties initiating sleep^2^. Many pharmaceutical treatments are available to treat insomnia. However, a recent meta-analysis found that many effective treatments were poorly tolerated, while well-tolerated drugs lacked evidence supporting their efficacy^3^. Recently, there has been growing interest in acoustic neuromodulation as an alternative to sleep medications for improving sleep quality. This approach uses sound pulses phase-locked to neural oscillations measured in real-time via electroencephalogram (EEG). Studies have shown this method to have health benefits when applied to slow (0.5-1.5 Hz) oscillations present during non-rapid eye movement (NREM) stage 3 (N3 or “deep”) sleep, including enhanced memory consolidation;^4–6^ immune function; ^7^ and autonomic balance.^8^ However, stimulation at slow-wave frequencies outside of N3 sleep has not been shown to improve sleep onset and in fact may delay the initiation of sleep.^9^ It remains unknown whether phase-locked acoustic stimulation can be used to address symptoms of insomnia.

Sleep can be described as cycling through four different phases, each defined by distinct patterns of neural activity as measured by EEG. Whereas N3 sleep is defined by the presence of high-amplitude oscillations in the 0.5 to 3.5 Hz range, the transition to sleep is often accompanied by high spectral power in the alpha (8-12 Hz) range while the sleeper is awake with closed eyes. The shift from wake to phase N1 sleep is defined in part by a loss of this alpha power.^10^ Interestingly, the strength of alpha oscillations has been shown to negatively correlate with feelings of sleepiness^11^ and sleep depth,^12^ and alpha power during sleep is known to be elevated in insomnia.^13,14^ Therefore, disruption of this alpha process represents a potential target to promote sleep.

Although the role of alpha oscillations in sleep onset is not fully understood, alpha more broadly has been shown to regulate sensory processing and attention in a phase-specific manner^15,16^ with alpha peak phases corresponding to inhibited neural activity states and trough phases associated with excitable states^17^. Alpha phase can influence the neural response to sensory stimuli^18,19^ as well as the conscious perception of sensory events^20^. Reciprocally, evoked responses generated by sensory events have been shown to impact ongoing alpha oscillations. The power of alpha following a sensory evoked response is modulated differently depending on the phase of alpha at which it was delivered^21–23^, and alteration of alpha oscillations by sensory or electrical stimulation has direct consequences on perception and cognition^24–26^. Given the association between alpha power and heightened arousal before and during sleep, we asked whether the neuromodulatory effects of alpha-phase-specific sensory stimulation may have an impact on sleep onset. We hypothesized that aligning an auditory evoked response potential (ERP) with the excitable trough phase of alpha, an approach which has been shown to decrease subsequent alpha power^18,21,27,28^, may hasten the transition from wakefulness to sleep in individuals experiencing insomnia symptoms. Such an approach has the potential to extend the impact of closed-loop acoustic neuromodulation beyond N3 sleep and provide an alternative to sleep medications for accelerating sleep onset.

To answer this question, we conducted a multi-night, randomized controlled crossover trial using a wearable system, the Elemind Neuromodulation device (ENMod). The ENMod consisted of a wearable headband with 3 EEG channels roughly corresponding to Fp1, Fp2, and Fpz according to the 10-20 convention and was controlled by a custom smartphone application. Computation of instantaneous alpha phase was accomplished using an endpoint-corrected Hilbert transform^29^ implemented on the device. During the Stimulation week, the system was programmed to deliver alpha phase-locked auditory stimulation for 30 minutes at the beginning of the night. During the Sham week, the headband silently recorded EEG. The primary objective of the study was to evaluate the effect of phase-locked auditory stimulation on SOL for individuals with objectively verified sleep onset insomnia symptoms.

## Methods

### Study Design

A single-arm, randomized controlled crossover design was used which consisted of a 2-night sham run-in (Week 0, Thursday and Friday), during which participants acclimated to use of the headband and were screened for compliance with study procedures as well as prolonged sleep onset latencies (SOL > 30 minutes, assessed by EEG). Following randomization, participants underwent 4 nights of sham or active stimulation (Week 1, Monday through Thursday), a washout period (Week 1 Friday through Sunday), and finally 4 nights of the remaining condition (Week 2, Monday through Thursday) (Figure 1). Participants completed the study in their own homes. After enrollment but prior to beginning the study, participants were trained to operate study equipment over video call. This included use of the companion smartphone app to start/stop sessions as well as to ensure good signal quality from the EEG electrodes. Additionally, participants were instructed on the study protocol, and asked to maintain a consistent bedtime routine and sleep schedule for the duration of the trial. Studies were approved by an independent institutional review board (Solutions IRB, Yarnell, AZ). All experiments were performed in accordance with relevant guidelines and regulations. The trial was registered on clinicaltrials.gov under registry number NCT05743114. The date of first registration for the trial was 24/02/2023.

**Figure 1:**
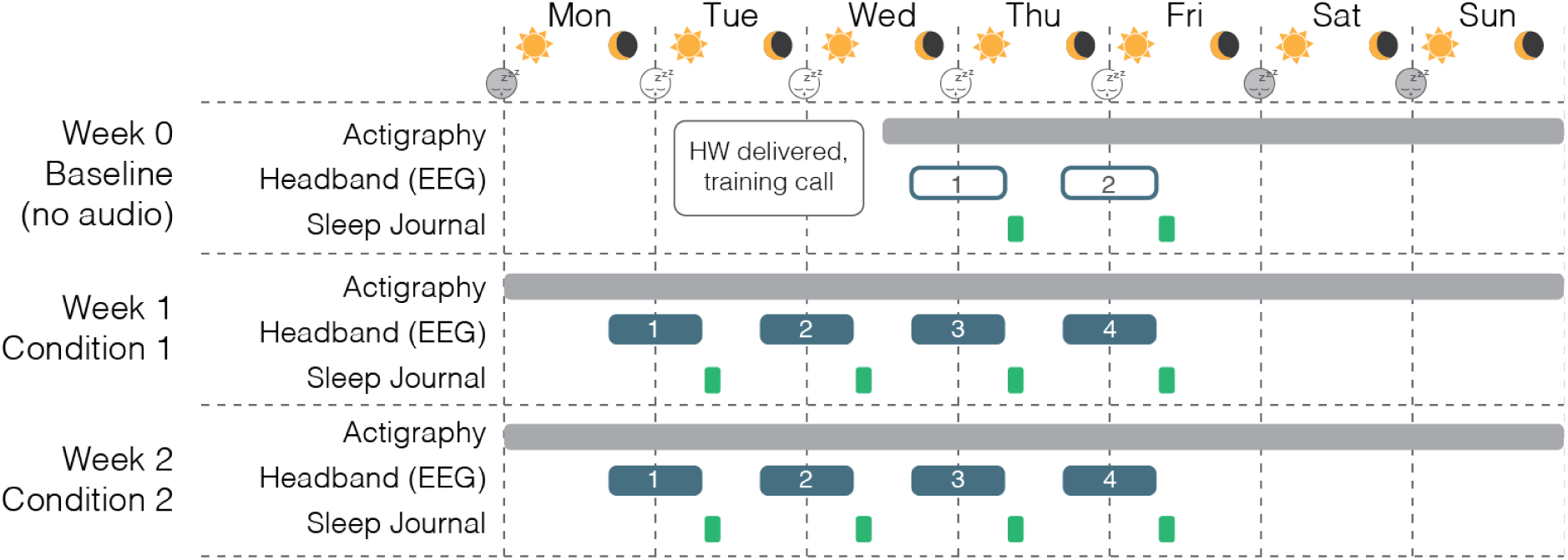
Design of the SLEEPFAST study. Baseline data collection for all subjects took place during Week 0, in which participants wore the headband with active EEG but without audio stimulation, and completed sleep surveys. Following randomization, four nights (Mon-Thu) of EEG and survey data were collected each week for 2 weeks of testing across 2 separate conditions: Sham (no audio), and Stimulation. Actigraphy was collected continuously for the duration of the study.

### Participants

Participants were recruited through online advertisements and eligibility was determined through an online screening questionnaire (Table 1) designed to identify individuals symptomatic of insomnia. Twenty-one participants including thirteen (13) females were included in the final analysis with an average age of 34.0 ± 6.1 years and a range of 22 to 44 years. All subjects reported typical sleep onset latencies of at least 30 minutes or more. Recruitment ended once the trial reached its enrollment target. Before conducting the study, written informed consent was obtained from all participants.

**Table 1:** Inclusion and exclusion criteria used to select participants.

| **Criteria** | **Inclusion** |
| --- | --- |
| **Language** | Fluency in English |
| **Internet access** | Access to internet or other cellular data service |
| **Insomnia Severity Index (ISI)** | Up to moderately severe but less than severe insomnia defined by an ISI score $\leq 21$ |
| **Pittsburgh Sleep Quality Index (PSQI)** | Poor sleep quality defined by a PSQI score $> 5$ |
| **Sleep apnea** | No clinically diagnosed sleep apnea |
| **Generalized Anxiety Disorder -7 (GAD-7)** | Up to moderate but less than severe anxiety as defined by a GAD-7 score $\leq 15$ |
| **Alcohol Use Disorders Identification Test-Concise (AUDIT-C)** | Less than high risk for alcohol use disorder according to an AUDIT-C score of $\leq 6$ |
| **Hearing** | No diagnosis of hearing impairment |
| **Body Mass Index (BMI)** | Not obese according to a BMI $\leq 33$ |
| **Night shift work** | No night shift work reported |
| **Pregnancy** | Not pregnant |
| **Medication use** | No reported antidepressant use, no hypo/hypertension medication, no cannabis use |
| **Stimulants** | No reported use of stimulants and $\leq 4$ caffeinated beverages per day |

### Randomization and Masking

Condition order for each participant was assigned using simple randomization via a computerized random number generator. Random order assignment was done after enrollment and the run-in period, and participants were not made aware of the condition order. Headbands were remotely programmed by research staff prior to the first night of each condition based on this random assignment. The ENMod headband was worn during both Stimulation and Sham conditions; however, due to the presence of sounds in the stimulation condition and lack of sounds in the sham condition, participants were not blinded to the intervention. During data log scoring, scorers were blinded to participant identity and condition associated with the log. A computer program was used to randomly select logs from a database and present them to the scorer.

### Procedures

#### Intervention

EEG phase-locked auditory stimulation was presented in the form of pink noise pulses delivered through a bone conduction driver positioned near the middle of the wearable headband (near the middle of the forehead). To create a more pleasant experience, a background of natural rain sounds were played at 18 dB below the volume of the pink noise. Volume for each session was set by the participants, who were instructed to adjust the volume of the rain sound such that it was just audible. Auditory stimulation began at lights-out and continued for 30 minutes.

The onset and offset of each pink noise pulse was gated by the current estimation of instantaneous alpha phase. Stimuli were presented at every cycle of alpha. Pink noise generation began when the phase estimation reached a predetermined onset phase, and ended when the phase estimate reached ¼ cycle (90°) after sound onset. Because there is a delay between sound presentation and elicitation of an auditory evoked response in the EEG, the onset phase was chosen to align the P1 component measured at electrode location Fpz of each pink noise-evoked response potential (ERP) to the trough of the next alpha oscillation cycle. This P1 delay was estimated to be 62.445 ms based on previous work^30^. Determining this onset phase also required estimating the peak of each subject’s individual alpha frequency (IAF, Supplementary Figure 1). IAF estimation was performed by identifying the spectral peak in the 7.5-12.5 Hz range following the first night of viable data collection during the run-in period.

#### Apparatus

Phase-locked auditory stimulation was delivered by the Elemind Neuromodulation Device (ENMod); a wearable electroencephalogram-enabled headband that computed the instantaneous phase of neural activity in the alpha (7.5 - 12.5 Hz) frequency band (see Supplementary Information) and delivered pink noise pulses at programmable onset and offset phases. The entire system consisted of 3 main components (Figure 2): 1) a compact processor unit (70 mm x 45 mm x 20 mm), weighing approximately 44 g including a rechargeable lithium-polymer battery and containing custom electronics used to collect EEG signals, perform phase estimation, and generate pink noise stimuli through a bone conduction driver. The electronics were operated via a Bluetooth LE connection and programmed over USB-C. This processor unit was mounted at the front of 2) a commercially available fabric headband (Muse-S Gen-2, InteraXon, Toronto, ON, Canada). The band included 6 flexible dry electrodes: 3 recording sites approximating Fp1, Fpz, and Fp2, a ground electrode adjacent to Fpz, and two linked reference electrodes positioned above the ears. Finally, participants operated the wearable device using 3) an Android smartphone (Samsung J3 Star, Samsung, Seoul, South Korea) running a custom application (Figure 2A). Participants also wore a wrist-worn activity tracker (Philips Respironics Actiwatch 2, Murrysville, PA) continuously from the first day of the study period until the conclusion of the study.

**Figure 2:**
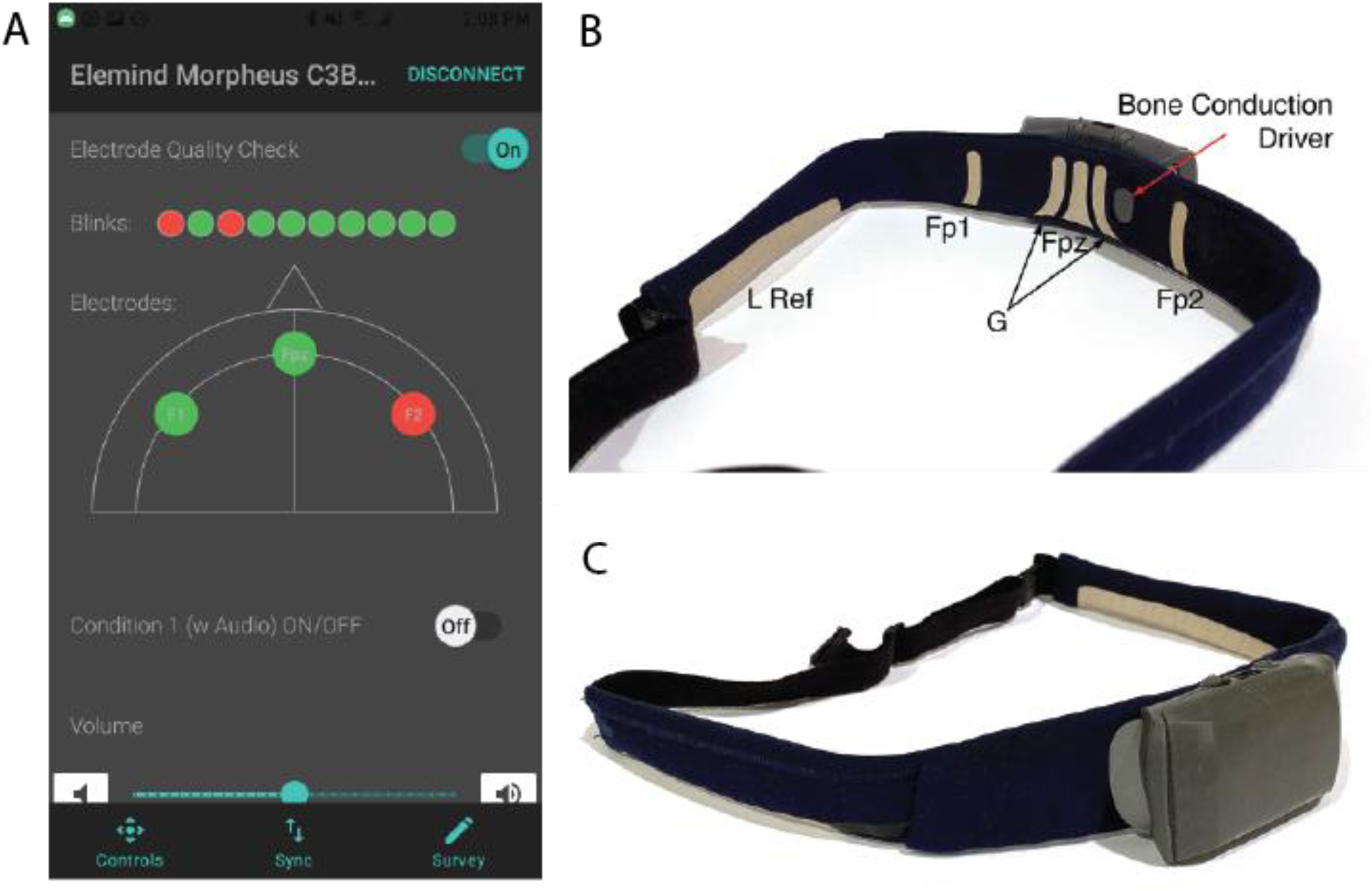
The Elemind Neuromodulation device (ENMod). (A) The primary UI of the smartphone app used to control the device. Users were given information about electrode contact using an eyeblink test and real-time signal quality of each frontal electrode.(B) Back view showing the left over-the-ear reference electrode (L Ref), recording electrodes Fp1, Fpz, and Fp2, ground electrode “G”, and the bone conduction driver. (C) Front ¾ view of the detachable processor unit attached to the fabric headband. Left over-the-ear reference electrode shown.

#### Data Collection

Actigraphy data from the Actiwatch was collected for the entire duration of the experiment period, including days and nights when the headband was not used. Participants used a button on the watch to manually mark bed and wake times. Actigraphy metrics were derived from the automatic scoring algorithms from the accompanying Philips Respironics Actiware software application (Version 6.2.0.39, Koninklijke Philips N.V., 2023).

Every morning following a night that the ENMod headband was used, participants filled out a sleep survey (Supplemental Table 2). The survey contained questions asking about the previous night’s sleep, including subjective sleep onset latency, total sleep duration, bedtime and wake time, subjective sleepiness upon waking, and questions relating to the experience of using the ENMod. EEG data was collected every night that the ENMod was worn, starting from lights-out and continuing for 2 hours (10 nights total for each participant). EEG data was sampled from each of the 3 channels (Fp1, Fpz, and Fp2) at 250 Hz and 24-bit resolution, and bandpass filtered online with a lowpass cutoff of 35 Hz and highpass cutoff of 2.5 Hz. Signals were referenced to a linked pair of reference electrodes located at the exposed skin regions just behind the helix of the pinnae. The ground electrode was located adjacent to electrode Fpz.

#### Sleep Stage Scoring

Data logs from the ENMod and morning survey results were screened by the research team each morning following a night of data collection. During the sham run-in period, participants who did not comply with the study protocol, or who exhibited EEG-verified sleep onset times of 30 minutes or less were excluded from further participation. EEG-based sleep staging analysis was done by two visual scorers working independently. Scorers were blind to subject identity and experimental conditions of each data log. Scorers reviewed each 2-hour data log in 30-second epochs using a custom data visualization program (Supplementary Figure 4), and marked the onset time of the first observable sleep spindle. Results from each scorer were compared using a Bland-Altman analysis (Supplemental Figure 3). Data logs were flagged if scorers disagreed on scorability based on data quality, or if the scored sleep onset time fell outside of the 95% confidence interval of the bias estimate. Flagged data logs were re-blinded and reanalyzed by both scorers working together to reach consensus. Not all EEG logs were of sufficient quality for determining sleep onset. Data logs in which data loss or low data quality occurred at critical sleep stage transitions were excluded from analysis. These inclusion/exclusion decisions were made by the visual scorer without knowledge of the participant or experimental condition of the log. All actigraphy-based analyses including sleep onset latency (SOL) were derived from the automatic scoring algorithms using the Philips Respironics Actiware software application that accompanies the Actiwatch devices worn by participants (Version 6.2.0.39, Koninklijke Philips N.V., 2023)

### Outcomes

The outcome measures included comparisons between both treatment conditions (Stimulation and Sham) for each subject. The primary outcome measure was sleep onset latency (SOL) determined by blinded scoring of EEG records. SOL was defined as the elapsed time between lights out and the first identifiable sleep spindle, a hallmark of NREM sleep. Secondary outcome measures included the standard deviation of SOL (variability), and several sleep metrics measured using an automated actigraph algorithm as well as subjective post-sleep surveys. Actigraph-based metrics included Total Sleep Time (TST), Sleep Onset Latency (SOL), Sleep Efficiency (Eff), Wake After Sleep Onset duration (WASO), and number of WASO events (nWASO). Subjective measures included SOL, TST, WASO, and overall impression of sleep quality. We also measured EEG-based outcomes including the number of spindles, spindle power, spindle frequency, and alpha frequency variability (AFV).

### Statistical Analysis

#### Sample Size Determination

A power analysis was performed to determine the target study population size. As confirmed in multiple clinical studies, subjects with clinically confirmed insomnia disorder have EEG-based sleep onset to stage-2 (SOL-N2) ranging from 13.40 to 57.80 minutes.^13^ This study selected subjects using criteria similar to these prior studies and a pilot feasibility study^30^ which included 7 subjects. In our feasibility study, the resulting group mean sleep onset latency was about 35 minutes (SOL-N2 = 35.30 ± 13 minutes) for the Sham “No Audio” condition. Based on this SOL-N2 mean and standard deviation of 35.30 ± 13, a two-tailed t-test analysis assuming a type I error rate of α = 0.01 at 90% power required 15 subjects and 21 subjects to confirm a 15 and 12 minute reduction in SOL, respectively.

#### Statistical Model

Multilevel model statistical comparison testing was performed in R using the lme4 library to determine which fixed and random effects had a significant effect on sleep onset latency as determined by the time to first identifiable spindle. To determine which random effects terms to include, we constructed a maximal model that included Condition (Sham or Stimulation), Week (first or second week following the sham run-in period), Order (sequential order of data collection night in each condition, 1-4), and Weeknight (categorical: Sun-Fri, no data collection on Sat) as fixed effects; the random fixed effects structure included random intercepts for Subject and a random slope term for Condition for each Subject (Eq. 1).

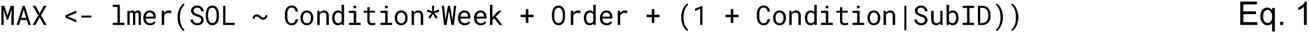

Three reduced sub-models were tested against the maximal model, each systematically modifying a single random effects term. The first model, RE1, removed the correlation between the random intercept of Subject from the random slope of Condition (Eq. 2). The second, RE2, modeled both Condition and Subject as independent random intercepts (Eq. 3). The third model, RE3, only considered Subject as a random intercept (Eq. 4).

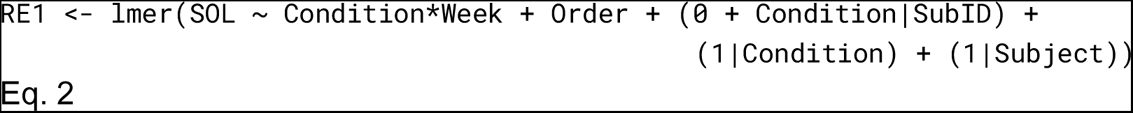

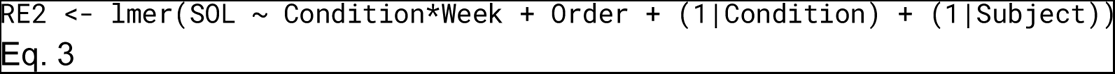

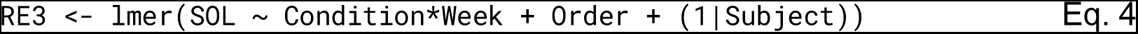

Model RE3 (Eq. 4) was ultimately selected based on likelihood ratio testing (LRT) and the resulting Akaike (AIC) and Bayesian (BIC) Information Criteria. This structure was used as the maximal model for evaluating which fixed effects to include in the final linear mixed effects regression model for each all sleep outcome measures, primary and secondary.

Final selection of the most parsimonious fixed effects structure was also performed using the LRT, AIC, and BIC selection criteria.

One-tailed paired t tests were used to compare weekly-averaged sleep outcome measures across Condition (Sham vs. Stimulation).

### Results

#### Trial Population

Between September 1st, 2021 through December 15th, 2022, 450 patients were assessed for eligibility, of which 155 met the inclusion criteria. 100 total patients chose to enroll in the study following the information and consent call and received an ENMod headband. Figure 3 shows the CONSORT flow chart. During the sham run-in (Week 1), 35 subjects were disqualified when they did not exhibit sleep onset latencies of greater than 30 minutes as scored by EEG, or if they did not comply with the study protocol (See Supplementary Table 1 for characteristics of these patients). 3 participants were dismissed due to skin irritation from the headband, and 8 participants withdrew for other reasons before completing the study. 54 participants completed all 3 weeks of the trial. During blinded scoring of EEG recordings, data from 30 participants had to be excluded from analysis for not meeting quality criteria, and a further 3 participants were excluded when none of their data logs in either condition displayed sleep onsets greater than 30 minutes (good sleepers). The remaining 21 subjects were included for the final analysis. Due to a technical failure in one of the Actiwatches, the analysis of actigraphy results contains data from 20 of the 21 subjects.

**Figure 3:**
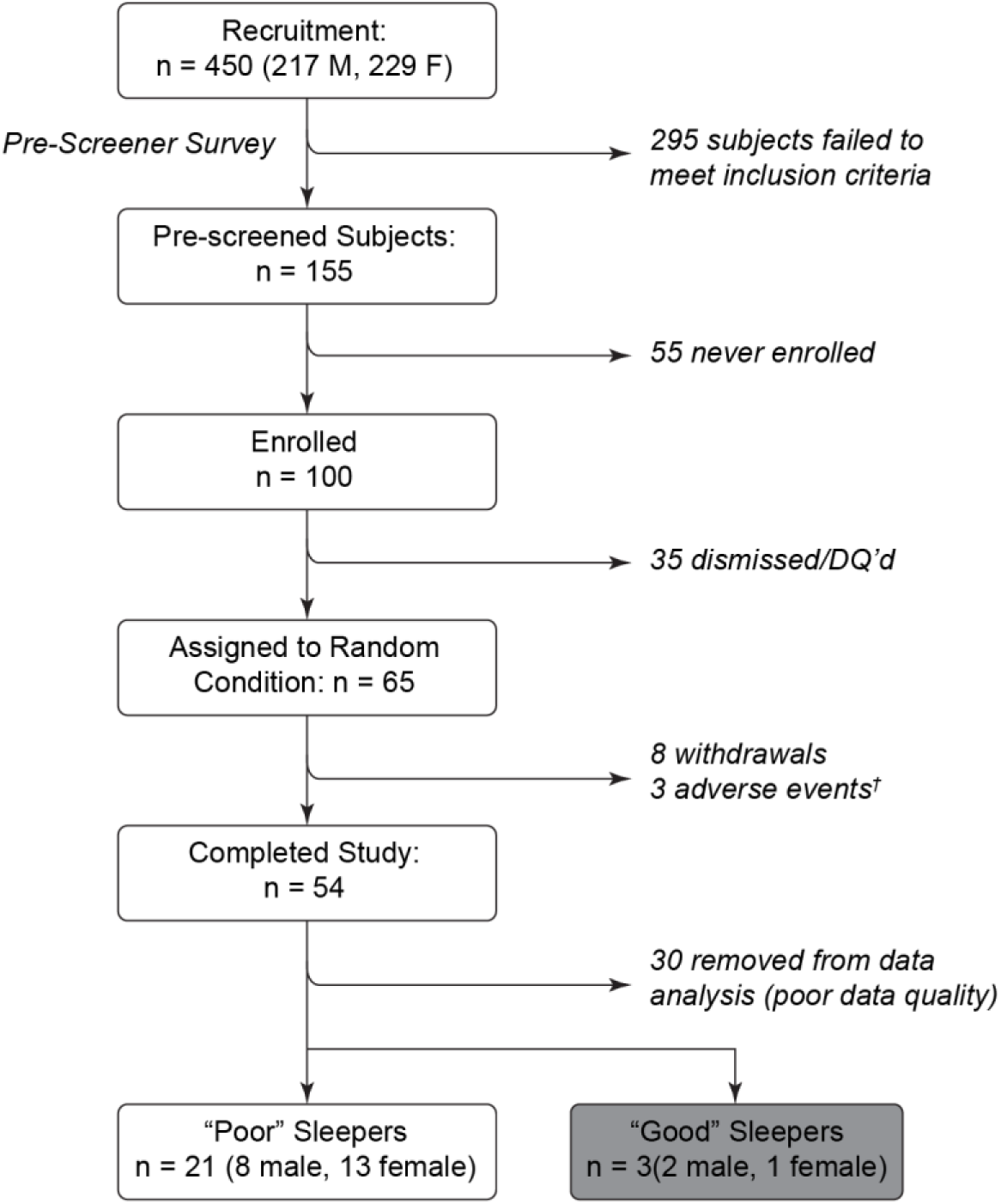
CONSORT diagram detailing the flow of participants throughout the study. ^✝^All adverse events were related to mild skin irritation from skin contact with the bone conduction driver.

#### Baseline characteristics

Table 2 lists the baseline characteristics of the subjects collected during the screening process as well as the run-in period.13 (61.9%) of analyzed participants were female. The average age was 34 ± 6.1 years with a range of [22,44] years. All participants were classified as Poor Sleepers according to the Pittsburgh Sleep Quality Index (PSQI), with scores > 5. The mean PSQI score was 11.0 with a standard deviation of 2.0. The Insomnia Severity Index (ISI) scores reflected no insomnia to moderate insomnia, with most participants being classified as Sub-Threshold (n=7) and Moderate (n=10). The four remaining participants fell into the No Insomnia category. Reported time to fall asleep on a typical night was 30 minutes (n=9), 45 minutes (n=5), 1 hour (n=5), 1.5 hours (n=1) 2 hours (n=0) and >2 hours (n=1).

**Table 2:** Baseline characteristics of included study participants. BMI: Body Mass Index; SOL: Sleep Onset Latency; TST: Total Sleep Time; Eff: Sleep efficiency; WASO: Wake After Sleep Onset, nWASO: Number of WASO events, PSQI: Pittsburgh Sleep Quality Index, ISI: Insomnia Severity Index, EEG: Electroencephalogram

| Variable | Mean ± SD |
| --- | --- |
| Age | 34 ± 6.1 |
| Gender (female) n (%) | 13 (61.9%) |
| Body Mass Index (BMI) | 24.5 ± 3.8 |
| Reported time to fall asleep on a typical night (minutes) | 42.2 ± 18.1 |
| Number of caffeinated beverages per day | 0.7 ± 0.8 |
| Pittsburgh Sleep Quality Index (PSQI) score | 11.0 ± 2.0 |
| Insomnia Severity Index (ISI) score | 13.0 ± 4.7 |
| EEG estimated Sleep Onset Latency (SOL) (minutes) | 27.7 ± 15.4 |
| Actigraphy estimated SOL (minutes) | 19.2 ± 17.1 |
| Actigraphy estimated Total Sleep Time (TST) (hours) | 5.8 ± 1.3 |
| Actigraphy estimated Sleep Efficiency (Eff) | 82.4 ± 8.3 |
| Actigraphy estimated Wake After Sleep Onset (WASO) (minutes) | 42.5 ± 23.4 |
| Actigraphy estimated number of WASO events (nWASO) | 20.9 ± 9.4 |
| Subjective SOL (minutes) | 40.6 ± 16.3 |
| Subjective TST (hours) | 6.4 ± 2.1 |
| Subjective WASO (minutes) | 16.3 ± 15.5 |
| Subjective Sleep Quality (0-4 scale) | 1.96 ± 0.6 |

#### Device performance

Across all 30-minute stimulation sessions, the mean number of auditory stimulation pulses delivered per session was 15341.78, with a standard error of 214.46 pulses. The mean pulse duration per session was 28.00 ms, with a standard error of 0.01 ms. To determine that the device correctly delivered auditory stimulation at the intended target phase during the at-home study, we computed the phase accuracy of the stimulation pulses using a post-hoc analysis of recorded EEG activity and stimulation timestamps. At every cycle of alpha, stimulation was programmed to begin at a specific target onset phase, and end at a target offset phase 90 degrees later. Phase onsets and offsets were programmed for each subject according to their individual alpha center frequency (IAF) to align the auditory evoked response anti-phase to the next alpha cycle (See Methods and Supplementary Data). We therefore computed the accuracy of both onset and offset timing for each stimulation pulse separately. Phase error (the difference between target phase and the actual phase that a pulse started or ended) was on average 0.5° ± 6.9° for pulse onsets, and 0.6° ± 14.1° for pulse offsets (Figure 4A, B). To measure the consistency of stimulation timing relative to alpha phase, we computed the phase locking value (PLV) for each session. The PLV is a measure of variability, such that a PLV value of 1 would indicate all pulses arrived at exactly the same phase, while a PLV near 0 would indicate that pulses arrived at randomly distributed phases ^31^. Across all sessions, the median PLV was 0.59004 for pulse onsets and 0.32527 for pulse offsets (Figure 4C; Supplementary Figure 5).

**Figure 4:**
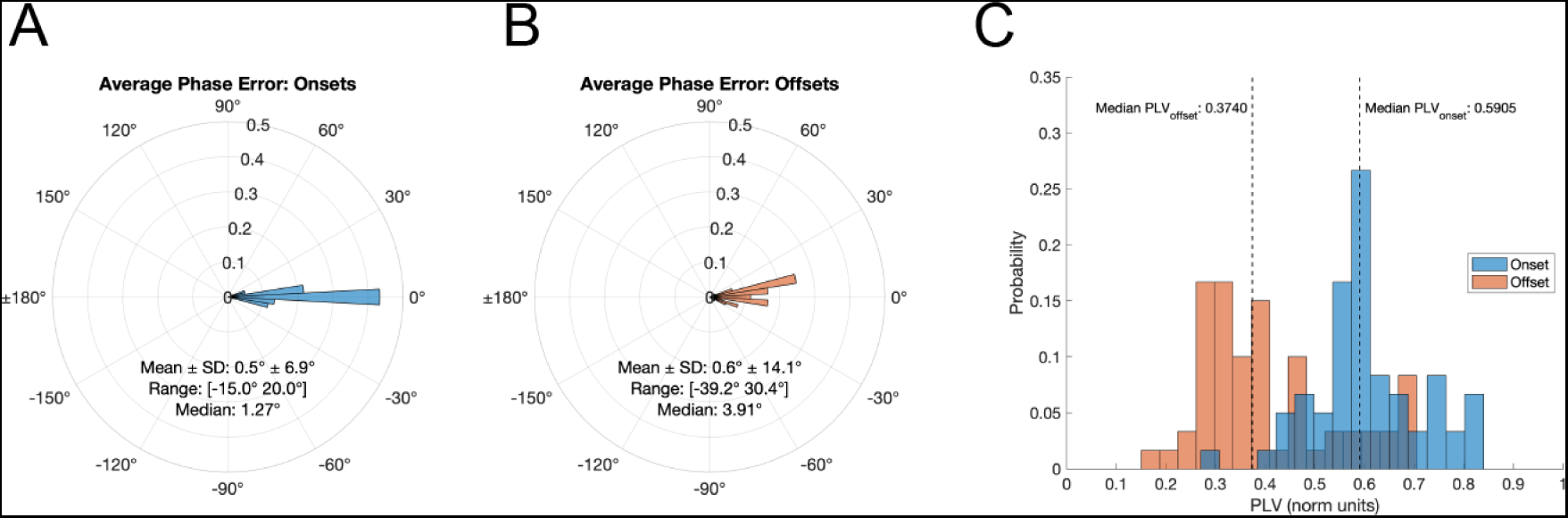
Summary of the phase locking accuracy of the pink noise pulse onsets (blue) and offsets (red) to alpha activity. A and B: Polar probability histograms and summary circular statistics of the mean phase error for pulse onsets (A) and offsets (B) for all available data logs (n = 60). C: Probability distributions of the per-data log phase locking values for all available data logs. Median PLVs are delineated by the dashed vertical lines.

#### SOL outcome measures

Sleep onset was defined as the time elapsed between lights-out and identification of the first sleep spindle, an EEG feature that is a hallmark of NREM sleep. A linear mixed effects regression analysis found a significant main effect of stimulus condition on sleep onset latency as measured by the time to the first identified sleep spindle [F(1,114.28) = 10.08, p = 0.001927]. Overall, the size of this improvement on sleep onset latency was 9.62 minutes (9 min 37 sec) based on the estimated marginal means of 35.1 minutes for Sham and 25.5 minutes for Stimulation (Figure 5A). Other fixed effects considered for analysis (recording night (1st vs 2nd vs 3rd vs 4th), study week (Week 1 vs Week 2), and Weeknight (Mon vs Tue vs Wed vs Thu)) were not found to be statistically significant predictors of sleep onset latency (Table 3).

**Figure 5:**
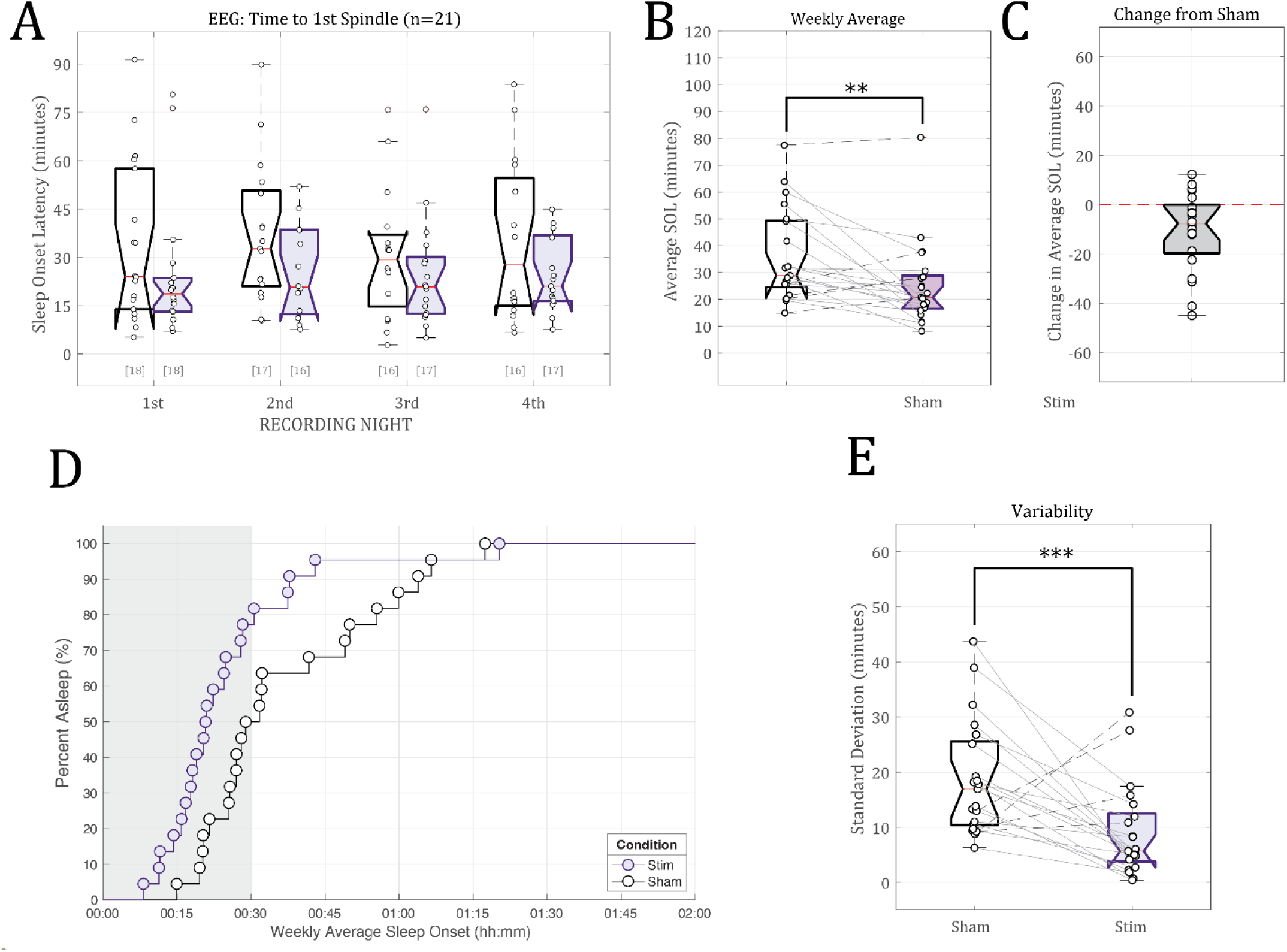
Sleep onset as defined by the time (in minutes) of the first visually-identified spindle in the EEG data. A) Individual subject time to 1st spindle estimates for the Sham control condition (white) and the phase-locked auditory stimulation condition (purple). The bracketed numbers below show the number of valid data logs (out of 21 subjects). B) The across-night weekly average latency to 1st spindle. Individual subject trends across conditions are plotted using connecting lines between data points. Subjects whose average sleep onset latency decreased are connected with solid lines; subjects whose average weekly sleep onset times increased are connected with dashed lines. C) Per-subject difference from Sham (no stimulation) weekly sleep onset latency estimates due to phase-locked stimulation (Stim - Sham). Subjects with faster weekly sleep onset times have negative values. D) Survivor plot showing the weekly average time to fall asleep for all participants in the Stim (purple circles) or Sham (white circles) conditions. The shaded gray bar represents the 30-minute stimulation period (Stim sessions only). E) Weekly standard deviation in subjects’ sleep onset times (in minutes). Standard deviations in sleep onset times were calculated from available data from the four nights of recorded EEG data for each of the two conditions** indicates p<0.05, *** indicates p<0.001.

**Table 3:**
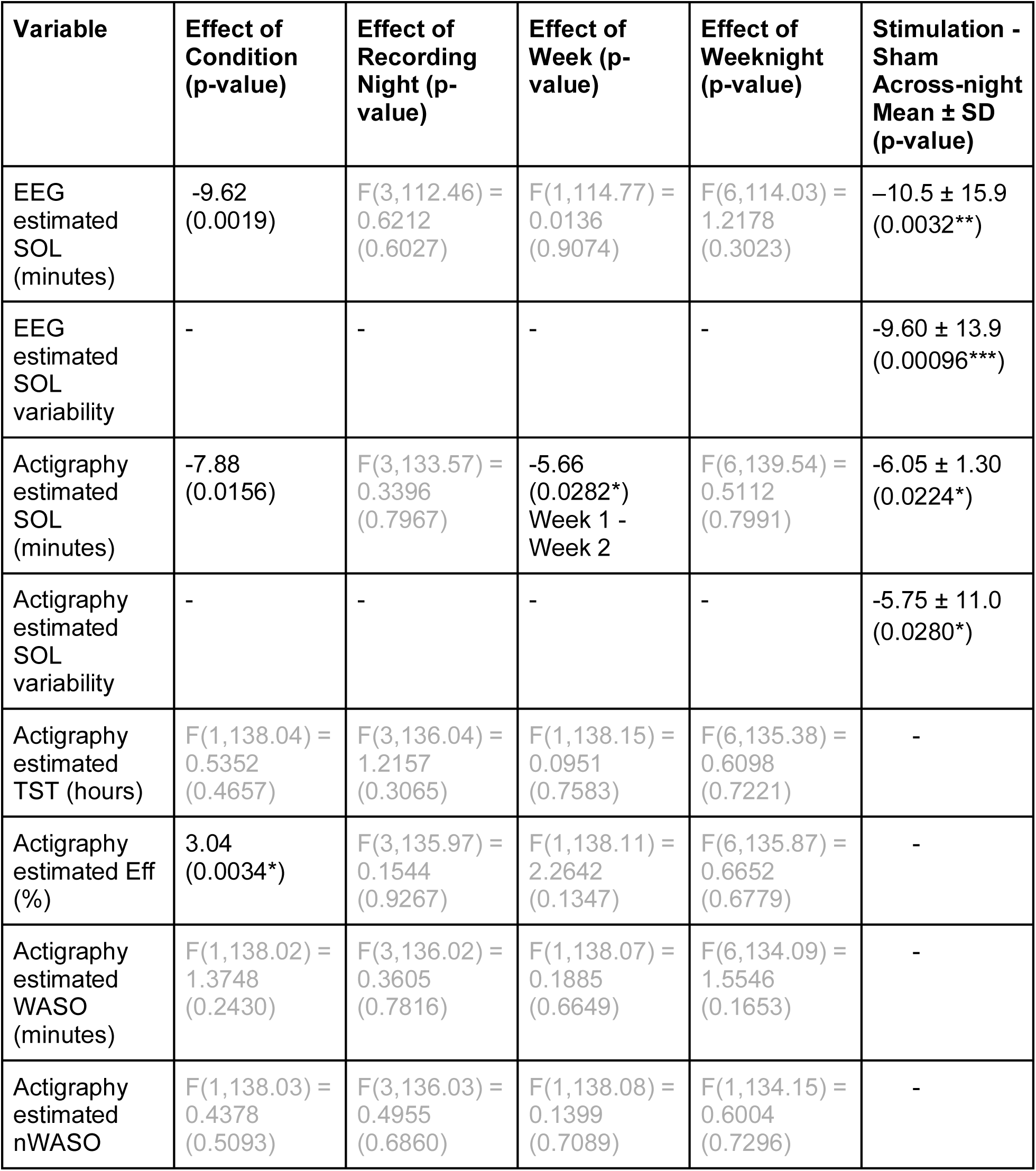
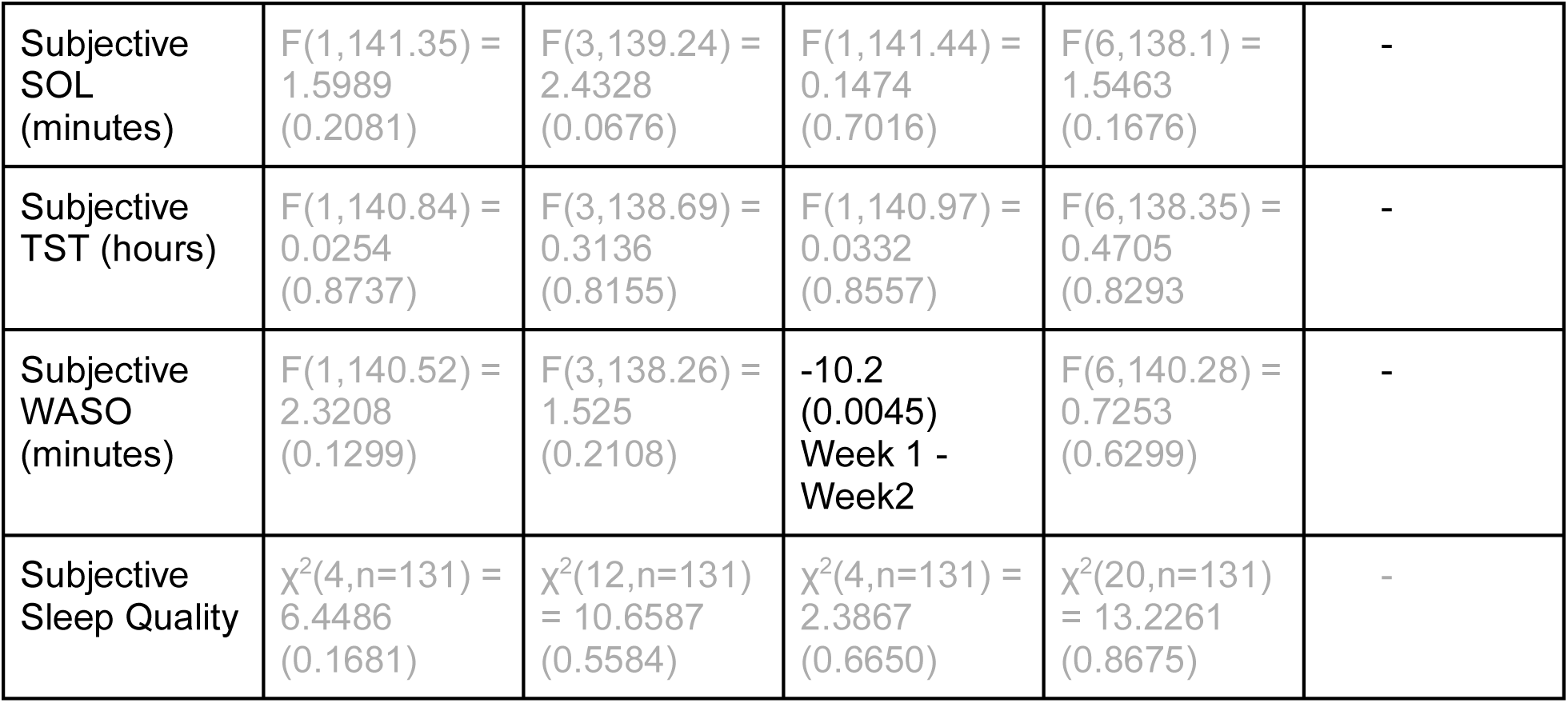
Primary and secondary outcome metrics.

Time to fall asleep can vary from night to night based on a number of life circumstances. To gain a holistic understanding of how stimulation affected sleep onset latency for each participant during the testing weeks, we quantified the weekly means and standard deviations for EEG-estimated SOL for both the sham and stimulation conditions. We observed a statistically significant drop in weekly average SOL from an average of 35.8 ± 17.0 minutes during the sham week versus 25.3 ± 15.6 minutes in the stimulation condition, or a 10.5 ± 15.9 minute average improvement in sleep onset times [One-tailed t test: t(20) = 3.0417, p = 0.0032] (Figures 5B and 5C). On average, 81% of participants fell asleep during the 30-minute Stimulation period compared to only 50% during the first 30 minutes of Sham nights (Figure 5D, Kaplan-Meier log-rank test: p = 0.0018, chi-squared(df = 1) = 9.7009). Furthermore, the consistency of sleep onset time was also significantly different between the two testing weeks. The weekly standard deviation in sleep onset times was reduced by 9.60 ± 13.9 minutes during the week that participants received active stimulation compared to sham (18.6 ± 10.4 minutes for sham vs 9.02 ± 8.26 minutes for stim [one-tailed t test: t(40) = 3.3214, p = 0.00096], Figure 5E).

A similar effect of stimulus condition was corroborated by actigraphy-estimated sleep onset latencies using a separate linear mixed-effects regression analysis on actigraphy data [F(1,135.661) = 5.4942, p = 0.02053]. Although sleep onset times here were earlier than what we observed using the sleep spindle metric, we still observed later sleep onset times during the control sessions (17.6 minutes) compared to phase-locked stimulation sleep onset times (11.6 minutes, Figure 6A). The total estimated improvement, as calculated by the estimated marginal means, was 5.98 minutes (5 min 59 sec). Unlike EEG metrics, we additionally observed a statistically significant main effect of “week” (Study Week 1 vs Week 2), where recorded sleep onset times were later in Week 2 compared to Week 1, regardless of the assigned condition [F(1,136.033) = 3.9400, p = 0.04916]. The estimated marginal means estimated this difference to be 5.62 minutes (12.1 minutes for Week 1 vs 17.2 minutes for Week 2). No interaction between “condition” and “week” was found. Weekly average SOLs also differed significantly according to actigraphy. The average sleep onset time for the control condition was 17.0 ± 1.89 minutes versus 11.1 ± 1.89 minutes for the stimulation condition (Figure 6B). The average improvement in actigraphy-measured sleep onset times was 5.84 ± 13.3 minutes [One-tailed t-test: t(19) = 1.9603, p = 0.0324] (Figure 6C). Similar to EEG metrics, the standard deviation in SOL was also measured to be different between the two testing weeks using actigraphy: the reduction in weekly standard deviation was 5.75 ± 11.0 minutes; 15.3 ± 11.2 minutes for Sham vs 9.50 ± 6.53 minutes for Stimulation [one-tailed t test: t(39) = 1.9703, p = 0.0280 (Figure 6D)].

**Figure 6:**
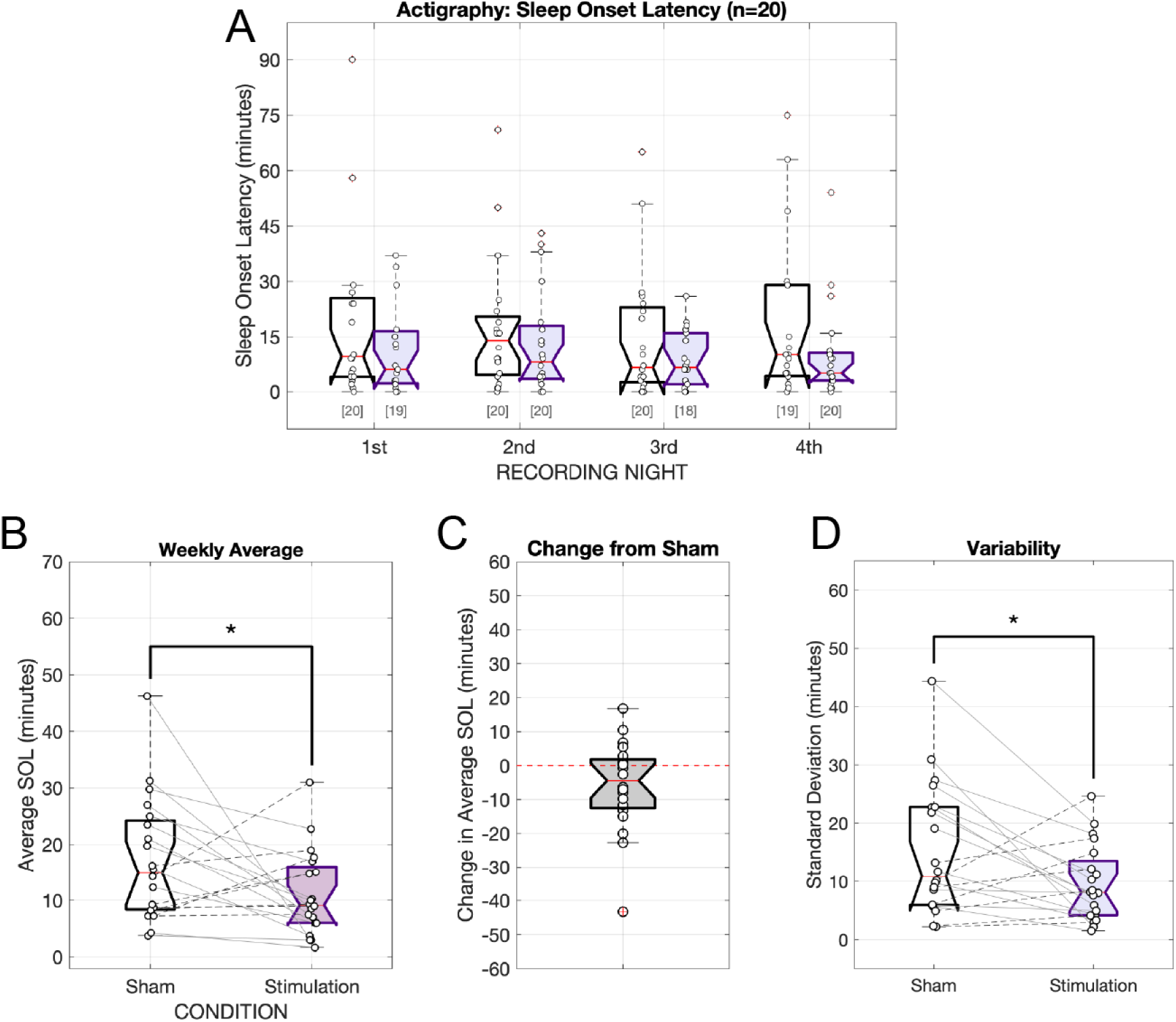
Sleep onset as estimated by wrist-worn activity trackers. A) Individual subject time to 1st spindle estimates for the Sham control condition (white) and the phase-locked auditory stimulation condition (purple). The bracketed numbers below show the number of valid data logs (out of 20 subjects). B) The across-night “weekly” average latency to 1st spindle. Individual subject trends across conditions are plotted using connecting lines between data points. Subjects whose average sleep onset latency decreased are connected with solid lines; subjects whose average weekly sleep onset times increased are connected with dashed lines. C) Per-subject difference from Sham (no stimulation) weekly sleep onset latency estimates due to stimulation (Stim - Sham). Subjects with faster weekly sleep onset times have negative values. D) Weekly standard deviation in subjects’ sleep onset times (in minutes). Standard deviations in sleep onset times were calculated from available data from the four nights of recorded EEG data for each of the two conditions. * indicates p<0.05, *** indicates p<0.001. Note: one (1) subject’s activity tracker suffered a technical failure and was not included in the actigraphy analysis.

#### Other Outcomes

For actigraphy-estimated sleep onset, The main effect of the stimulus condition was also reflected in the sleep efficiency scores [Condition: F(1,137.97= 8.8842, p = 0.003401]. Comparative analysis of the estimated marginal means found phase-locked auditory stimulation resulted in a small (3.04%) but significant increase in sleep efficiency (82.1% control/no audio vs. 85.1% stimulation). We found no significant effects of stimulus condition, recording night, study week, or weeknight on any of the other actigraphy-measured sleep outcome measures: total sleep time (TST), wake after sleep onset (WASO) duration, number of WASO events (nWASO) (Table 3). Survey-based metrics also did not differ between conditions (Table 4, Supplementary Table 3, 4, 5).

**Table 4:**
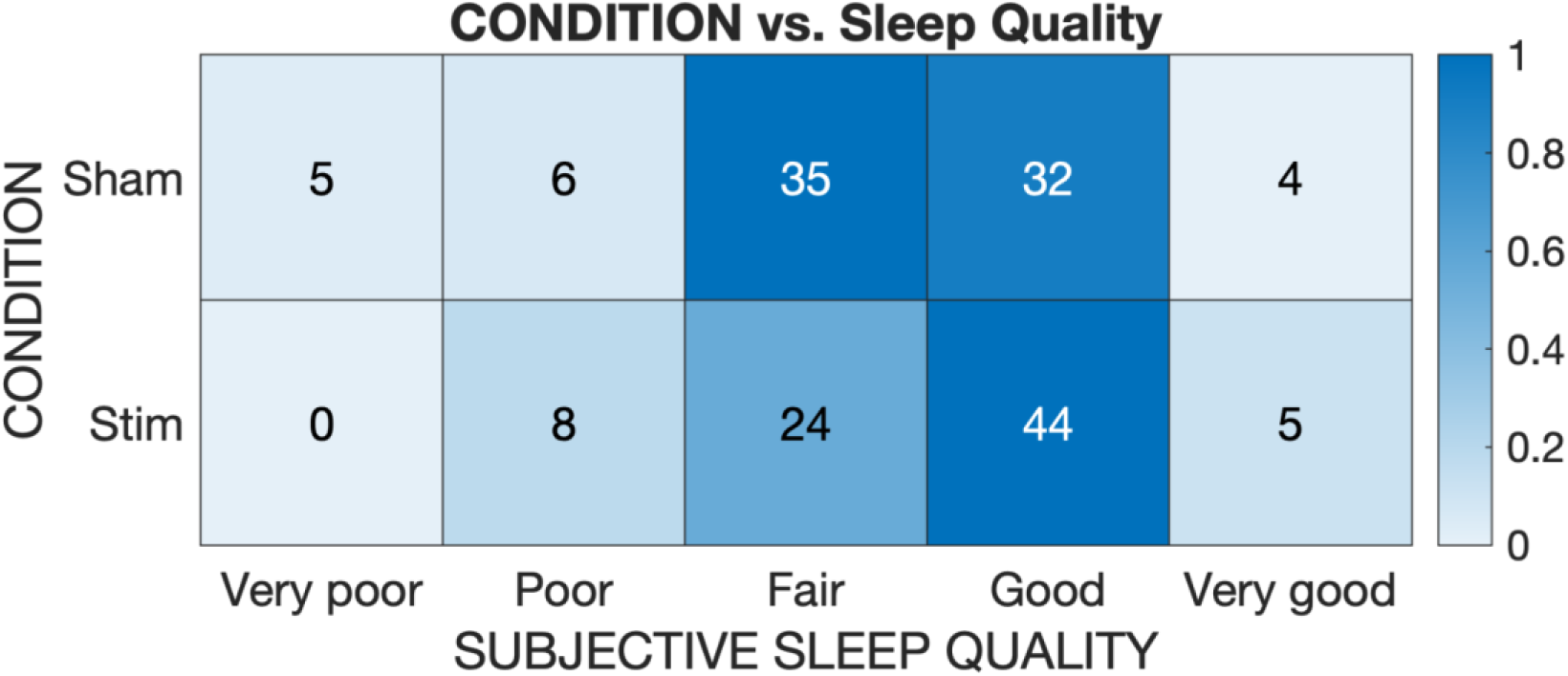
Heat map table showing the comparative number of responses (out of 5 possible) to a survey question asking “How would you rate the quality of your sleep?” Responses were collected each morning following use of the Elemind ENMod headband. Color intensity reflects the relative proportion of collected responses across rows. Total number of responses = 131.

We also examined other EEG-based features that may have been impacted by stimulation. Because our intervention targeted the alpha oscillation, we asked whether characteristics of alpha were altered in the Stimulation condition relative to Sham. Alpha Frequency Variability (AFV) is a metric that measures the variability across time in the instantaneous frequency of oscillations in the alpha band and has been shown to distinguish healthy sleepers and insomniacs^32,33^. Because insomniacs display a decrease in AFV relative to healthy sleepers specifically in the pre-sleep period^33^, we computed AFV in the 5-minute window at the start of each night, after participants had begun stimulation but prior to sleep onset. Pooling across all sessions, we found that AFV was significantly increased in this period during Stim sessions relative to Sham (Stim = 0.112 ± 0.047; Sham = 0.094 ± 0.033; P = 0.006, Figure 7A). These data suggest that alpha characteristics of our subjects more closely resembled healthy sleepers during stimulation.

**Figure 7:**
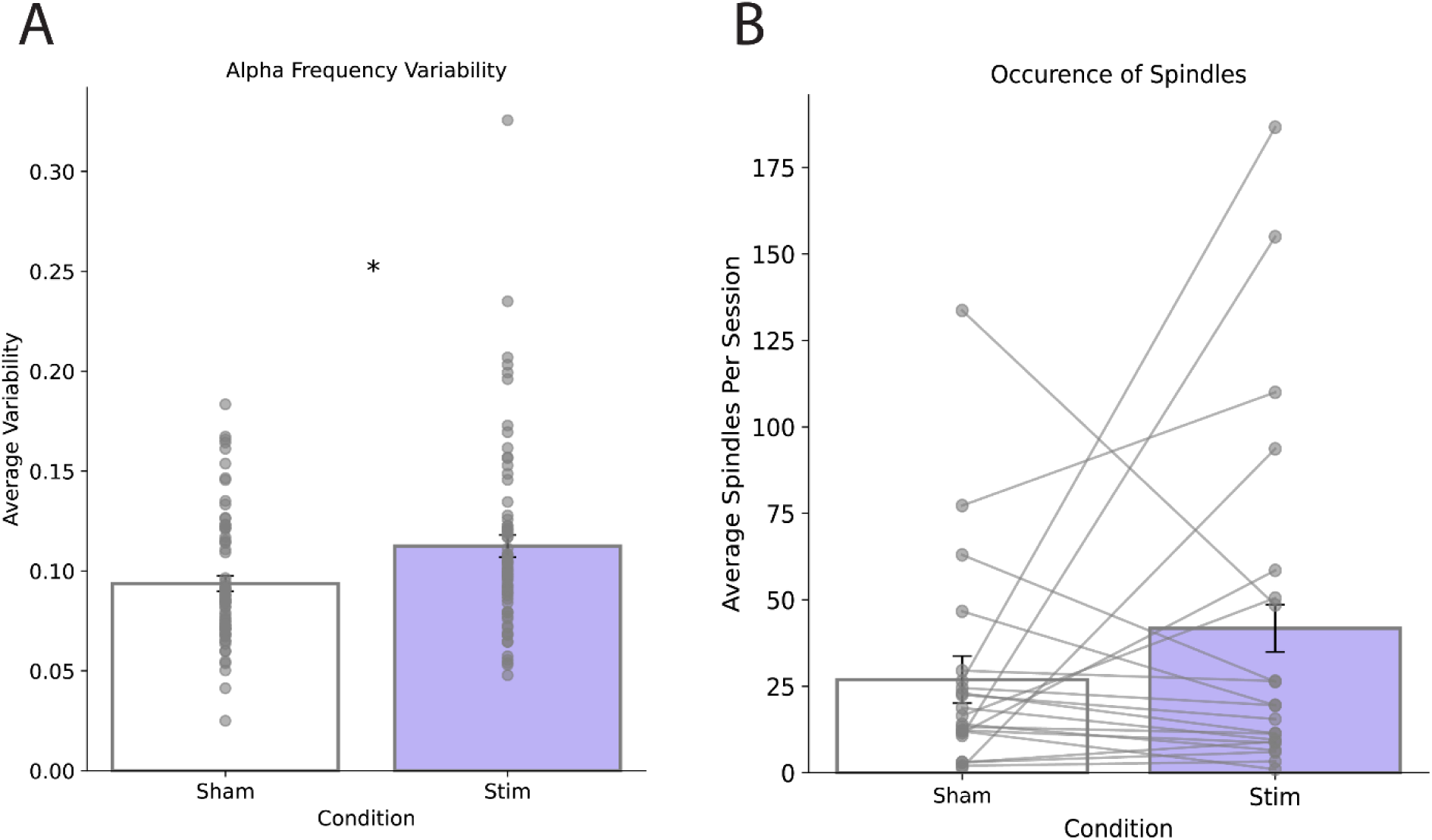
Other EEG-based outcomes. A) Change in alpha frequency variability (AFV) for Sham sessions compared to Stim sessions. AFV was computed over the first 5 minutes of each session. Asterisk denotes P<0.05. B) Occurrence of spindles during the first 30 minutes of all sessions between both conditions, averaged across participants. Lines connect points for each participant.

Because sleep spindles were readily apparent in our data, we also asked whether spindle characteristics were altered by stimulation. The total number of spindles during the first 30 minutes (the full stimulation period for the Stim condition) was slightly higher in the Stimulation condition (Sham = 25.73 ± 30.29; Stim = 39.92 ± 50.45; P = 0.244), but this difference was not significant (Figure 7B). Similarly, the average spindle duration and spindle power were not found to be different between conditions (Supplementary Figure 2).

#### Risk Ratio analysis

Following the standard clinical guidelines of diagnosing insomnia ^34^, we evaluated the total number of subject-nights where sleep onset (as defined by the time to the first sleep spindle) took longer than 30 minutes. For the sham condition, when no audio stimulation was given, over half of the subject-nights (50.8%) had sleep onset times of greater than 30 minutes. This rate dropped down to just over 25% when subjects were given phase-locked auditory stimulation. A relative risk ratio analysis shows that without auditory stimulation, subjects were nearly twice (relative risk ratio = 1.9717) as likely to experience a night where it took longer than 30 minutes to fall asleep compared to nights when subjects received active stimulation (Table 5). Additionally, using the absolute risk ratio of 0.2428, we can determine that the number needed to treat (NNT) is 4.119 nights; in other words, participants will on average have avoided a night of SOL > 30m after every 4.119 nights of use.

**Table 5:** Risk Ratios for sleep spindle-estimated sleep onset times.

|  | Sham | Stimulation |
| --- | --- | --- |
| # subject-nights SOL > 30 min | 34 | 18 |
| # total subject-nights | 67 | 68 |
| Event Rate | 0.5075 | 0.2647 |
| Absolute Risk Ratio (ARR) | – | 0.2428 |
| Relative Risk Ratio (RRR) | – | 1.9717 |

### Discussion

This is the first randomized controlled trial to test the application of acoustic stimulation phase-locked to alpha oscillations for the reduction of sleep onset insomnia symptoms. A significant decrease in SOL was observed in the Stimulation condition, with the majority (76.2%) of participants experiencing a benefit relative to sham. Only 3 of 55 subjects (5.4%) completing the study reported minor adverse events relating to skin irritation while wearing the headband (none of these subjects were part of the 21 included in the final analysis). Our results support the use of this approach to accelerate sleep onset for individuals with delayed sleep initiation.

Acoustic neuromodulation is a relatively new approach to augmenting sleep quality, and has primarily been investigated for the enhancement of slow-wave (N3) sleep. In these studies, delivering acoustic pulses at slow-wave frequencies has been shown to have neuromodulatory effects by influencing the power of subsequent oscillations as well as spindle activity.^7,9,35^ Notably, neuromodulatory effects appear to be dependent on phase-specific presentation of auditory stimuli,^5,36^ and similar phase-dependent effects have been also observed during the application of transcranial magnetic stimulation (TMS) during slow-wave sleep.^37^ These findings support the general notion that neural oscillations reflect states of fluctuating excitability, which can organize the spatiotemporal flow of information in the brain.^38,39^ Under this framework, it would be expected that the timing of stimulation relative to oscillatory phase would influence the magnitude or direction of the neuromodulatory effect.

Although the effect of phase-locked stimulation at alpha frequencies has not been previously investigated in the context of sleep, alpha phase-dependent effects have been observed in other domains. For example, transcranial magnetic stimulation (TMS) targeting subject-specific phases of alpha has shown promise in treating antidepressant-resistant major depressive disorder (MDD),^40,41^ possibly by engaging mechanisms of neuroplasticity. Manipulation of alpha phase by transcranial direct current stimulation (tDCS) has been shown to influence auditory perception thresholds,^24^ and transcranial alternating current (tACS) was used to manipulate crossmodal perception timing by influencing alpha peak frequency.^42^

Although there is a robust body of work describing the relevance of alpha oscillations in sensory perception and attention,^43^ the significance of alpha oscillations in early-stage sleep has yet to receive similar attention. Whether the role of alpha in sleep is related to or independent from its sensory and attention-binding features is unknown. However, intracranial recordings in humans have demonstrated a reduction in alpha desynchronization in response to auditory stimuli during sleep, which may be suggestive of a loss of feedback processing.^44^ Some evidence does exist to suggest an association between alpha and sleep initiation and maintenance. Aside from the loss of alpha power during the transition from wake to sleep, multiple studies have reported a positive correlation between the power of alpha oscillations during NREM sleep and insomnia symptoms.^13,45,46^ Additionally, a negative association between alpha power and sleep depth has also been observed.^12^ These findings may suggest a role for alpha power in the hyperarousal model of insomnia,^47^ but additional work is needed to better understand this link and the mechanisms behind it.

In our study, we set the timing of stimulation such that the auditory evoked response potential (ERP) of each acoustic stimulus arrived anti-phase to the alpha oscillation (i.e in time with the alpha trough, Supplementary Figure 1). Because auditory ERPs arrive after a sensory processing delay, this required customizing the trigger phase of the auditory onset and offset to align with each subject’s individual alpha peak frequency (IAF), based on a fixed ERP delay of ∼62 ms. This approach resembles stimulation protocols used to target alpha trough phase in MDD,^41^ but is a departure from SWS stimulation protocols that target the peak phase of slow waves. This choice was made based on the observation that slow wave power is positively correlated with desirable sleep outcomes, while alpha power is negatively correlated with sleepiness. However, the importance of this choice of target phase could not be determined from our data. It may also be the case that the ideal target phase is subject-dependent, which would require additional tuning. ^40^

Current sleep staging guidelines, which rely on a full complement of polysomnography sensors, have recommended scoring sleep onset as the time at which the first epoch can be classified as any stage of sleep.^10^ The addition of more sensors in our study (such as electromyogram, electrooculogram, and occipital channels) would have enabled a more thorough analysis of sleep macro- and microstructure and allowed for more traditional sleep scoring metrics to be used. However, our study design relied on participants administering the intervention independently in their own homes, and this would have been complicated by the need to also apply and operate a polysomnogram. Automated sleep staging algorithms have been developed as an alternative to manual sleep scoring and have demonstrated the capability to accurately score sleep with a limited number of input channels or even a single channel^48,49^. However, these algorithms show low performance when classifying stage N1 sleep, often identifying N1 epochs correctly less than 50% of the time^48,49^. Given these limitations, we chose instead to use the time of the first identifiable sleep spindle to determine sleep onset. Using time to first spindle as a marker of sleep onset is not without precedent,^50,51^ and may more closely match the subjective perception of sleep onset in insomniacs than other metrics.^52^ In fact, there is disagreement about when sleep truly begins, which likely occurs on a continuum rather than in a discrete moment. The occurrence of sleep spindles, however, is typically a sign that an individual is asleep.^53^ Future investigations, perhaps including overnight polysomnography or additional device features, could be conducted to better understand full-night effects on sleep structure.

The effect of therapy on our primary outcome measure (SOL) was also supported by actigraphy data, which is commonly used in at-home studies to measure sleep. Unlike EEG-based metrics however, a significant effect of Week was observed for actigraphy-based SOL, with subjects demonstrating a decrease in SOL during week 2 regardless of whether it was a Sham or Stim week. While accelerometer-based devices are often used to measure SOL and TST, they are often prone to error, likely due to the lack of neural data^54,55^. Because actigraphy algorithms use movement (or a lack thereof) to identify sleep onset, we see two possible explanations for this Week effect on SOL: 1) Subjects may have habituated to the study equipment over the course of participation, such that by Week 2, they were more quickly able to find a comfortable sleeping position or spent less time adjusting the headband. This would have reduced the amount of movement detected by the Actiwatch and led to an earlier SOL estimate. 2) Subjects may have benefited from learned improvements in sleep hygiene. Subjects were instructed to maintain a consistent sleep routine, including bed and wake times, and to refrain from using electronic devices after lights out. It is possible that by practicing these instructions, subjects experienced some accumulating benefit of reduced SOL outside of that which was related to the intervention. Indeed, these recommendations are commonly used in the behavioral treatment of insomnia, although evidence demonstrating their effectiveness is mixed^56^.

Our study excluded from analysis participants who did not present with at least one night of objectively verifiable SOL greater than 30 minutes. Although long SOLs were reported during pre-screening, this could only be verified in 65% (65 of 100) enrollees, and the remaining 35 were dismissed after the run-in period. There remains debate whether subjective or objective sleep metrics are more meaningful in regards to clinical outcomes; however, recent studies have demonstrated that self-reported sleep measures are prone to error and bias^57,58^, and do not correlate with daytime functioning^59^. Misperception of SOL is an especially common occurrence in insomnia, with recent studies suggesting a 163% difference between subjective and objective measures on average^60^. Sleep misperception may identify a unique insomnia subtype,^61^ or simply the fact that the boundary between wake and sleep is difficult to define objectively^53^. We observed a similar discrepancy in our included study participants, who did not report a significant change in subjective sleep onset during the stimulation week, despite the effects observed in actigraphy and EEG-based metrics. Regardless, our study was limited to participants with objectively delayed sleep onset, and therefore additional work will be needed to determine whether this approach can generalize to address a broader range of sleep onset insomnia complaints and whether there are additional effects on whole-night sleep structure.

Finally, limitations of the prototype ENMod device itself were revealed over the course of data collection that suggest improvements in future device designs. 30 subjects’ data was found to be of insufficient quality for accurate analysis, predominantly due to signal loss on the EEG channels. Redesign of the headband for better contact and the use of different electrode materials could help to mitigate this issue. Additionally, inclusion of more sensors and extending battery life would improve the quality and quantity of data collection and allow for full-night recordings. Expanding the sleep-tracking capabilities of this device will also require a redesign of the EEG acquisition hardware, which necessitated a high-pass filter setting of 2.5 Hz due to the presence of 2 Hz electronic noise. Improving the quality of the signal below this value will greatly enhance the ability of the system to detect slow wave activity which is characteristic of NREM Stage 3 sleep. Although pink noise has broad spectrum characteristics that make it a good choice for eliciting auditory ERPs, some subjects reported disliking the pink noise when trying to fall asleep. Creating different modulated sound designs may also be a way to improve performance and appeal to more users.

#### Conclusions

Delivering phase-locked auditory stimulation targeting alpha oscillations appears to be an effective intervention for treating sleep onset insomnia symptoms. Although the generalizability of the effect to broader groups of individuals has yet to be determined, this approach is effective at accelerating sleep onset on a scale similar to pharmaceuticals, and with fewer apparent negative side effects. Noninvasive, wearable systems for tracking sleep-related EEG and delivering closed-loop stimulation are promising tools to treat sleep disorders and promote healthy sleep.

## Data Availability

All data produced in the present study are available upon reasonable request to the authors

## Trial Registration

This trial was first registered on clinicaltrials.gov on 24/02/2023 under the name Sounds Locked to ElectroEncephalogram Phase For the Acceleration of Sleep Onset Time (SLEEPFAST), and assigned registry number NCT05743114.

## Acknowledgements

This work was funded by Elemind Technologies, Inc. The authors would like to thank Drs. Nir Grossman, Ed Boyden, Ruth Benca and Heather Read for input into experimental design and fruitful discussions.

## Author Contributions

Study concept and design: S.B., D.W. Device fabrication and programming: D.W. Data acquisition: S.B., R.Y. Data analysis: S.B., R.N., R.Y. Writing the manuscript: R.N., S.B. Revising the manuscript: S.B., R.N., R.Y. All authors had full access to all study data and had final responsibility for the decision to submit for publication.

## Data availability statement

Deidentified participant data and a corresponding data dictionary will be available with publication and upon request to the corresponding author. Elemind Technologies, Inc. will approve data sharing requests.

## Declaration of interest statement

Authors S.B., R.N., R.Y, and D.W. are employees of Elemind Technologies, Inc., which sponsored this study.

## Ethical considerations

This research was conducted in accordance with the principles of the declaration of Helsinki as well as local regulations. Studies were approved by an independent institutional review board (Solutions IRB, Yarnell, AZ). All experiments were performed in accordance with relevant guidelines and regulations. Before conducting the study, written informed consent was obtained from all participants.

## Notes

### Clinical Trial

NCT05743114

### Author Declarations

IRB of Solutions IRB gave ethical approval for this work

