## Supplemental Figures and Data for "A randomized controlled trial of alpha phase-locked auditory stimulation to treat symptoms of sleep onset insomnia"

**Supplementary Material**

Supplementary Methods

*Online computation of instantaneous phase.* The instantaneous phase of alpha-band EEG signals was performed online using an endpoint-corrected version of the Hilbert Transform (ecHT).[1] Similar to a standard Hilbert Transform, the procedure for the ecHT begins with transformation of a discrete time-domain signal $x_i[n]$ into the frequency domain using the fast Fourier transform (FFT). This signal, $X_i[f]$, is then transformed into the Hilbert analytic signal, $Y_i[f]$, by zeroing out negative frequency components and doubling positive frequency components. In a divergence from the standard Hilbert transform, the ecHT then multiplies the analytic signal by the frequency response of a causal 2nd-order Butterworth IIR bandpass filter, $H[f]$, with cutoff frequencies set to match the frequency range of interest (here, 7.5 Hz highpass and 12.5 Hz lowpass were used). Returning to the standard Hilbert workflow, the filtered analytic signal $Y_i[f]$ is then transformed back into the time domain using an inverse Fourier transform to yield the complex-valued analytic representation $z_i[n]$, containing a point-by-point estimate of the instantaneous phase and amplitude of the original signal $x_i[n]$. Unlike the standard Hilbert transform, the extra filtering step in the ecHT limits the oscillatory properties of the analytic signal to frequency components within the fitler's passband. This step corrects for the Gibbs phenomenon distortion due to non-uniform convergence of the discrete Fourier series at discontinuities between the beginning and end of the analytic signal. As a result, phase error at the end of the analytic signal (i.e. the most recent sample point) is minimized.

The ecHT was applied to EEG data over a 128-point sliding window that was incremented sample-by-sample. Phase was estimated based on the signal from a single active channel determined algorithmically to have the highest signal quality. To determine the active channel, the most recent 5 seconds of data was used to compute a root mean square (RMS) value for each of the three channels. If the RMS of the current active channel dropped below a fixed threshold of 2 μV, a new active channel was chosen to be the channel with the highest RMS value at that time.

*Computation of auditory stimulation phase timing.* Stimulation onset phases were programmed in order to align the P1 component of the auditory evoked response potential (ERP) such that it arrived anti-phase to the next cycle of alpha (Supplementary Figure 1A). This required knowledge of both the ERP delay relative to stimulus onset as well as the frequency of the alpha oscillation. The ERP delay was assumed to be 62.445 ms based on previous data measured from electrode location Fpz. Neural oscillations are non-stationary, meaning that they have a variable frequency. However, alpha is known to have an individualized fundamental center frequency (IAF) which is known be stable over time.[2] We estimated the IAF for each subject and used this value to program the stimulus onset phase. Stimulus offset phases were then programmed to be 90 degrees past the phase of onset.

*Estimation of IAF*: Using EEG data from the run-in nights, we computed the power spectral density of the time series data, and determined the IAF value by identifying the most prominent peak in the 8 - 12 Hz range. This peak can be best visualized by fitting a 3rd-order polynomial to the across-time median spectrum and subtracting the fit from the power spectral density plot to remove 1/f noise (Supplementary Figure 1b).

*Computation of stimulus onset phase to match alpha trough and peak:* Using the computed individual value for IAF and the fixed P1 delay, we computed the onset and offset phases as:

onset phase° = [-360° x P1 latency (sec) x IAF (Hz)] - 180°
offset phase° = onset phase° + 90°

The relationship between IAF, P1 latency, and stimulus onset/offset phases is shown in Supplementary Figure 1c and d.

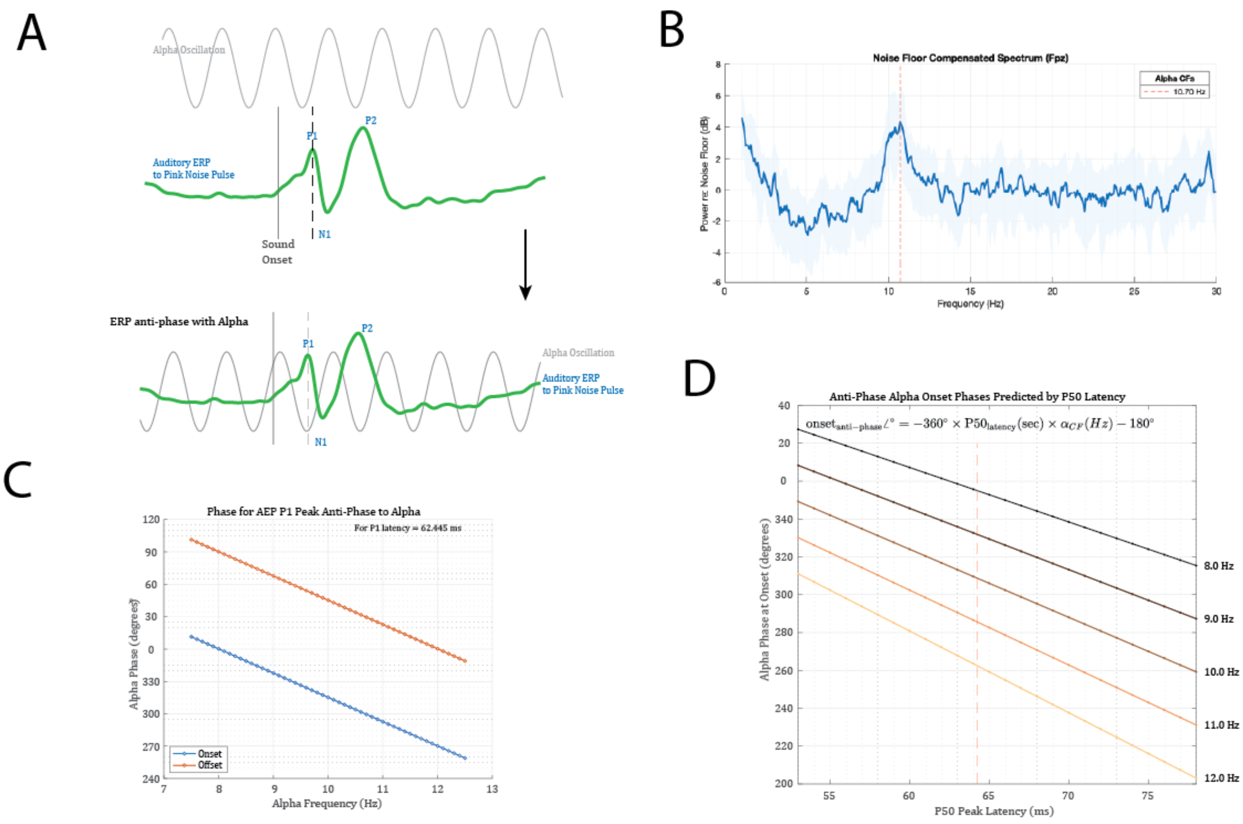

**Supplementary Figure 1:** Computation of stimulus onset and offset phases. A) The auditory evoked response potential (ERP) was aligned such that the P1 component of the ERP arrived anti-phase with the next alpha cycle. B) IAF was estimated for each individual from the power spectral density of EEG recordings made during the run-in period. C) Relationship between subject-specific individual alpha peak frequency (IAF) and programmed phase onset and offset values a function of frequency for the chosen fixed P50 value of 62.445 ms. D) Computation of onset phase, and the relationship between onset phase and ERP latency for several different IAF values.

*Determination of Alpha Frequency Variability*. Alpha frequency was determined using a bank of 6 length-3 IIR notch filters into which the bandpass-filtered (7.5-12.5 Hz) alpha signal was passed through[3]. The filters were evenly spaced at 1-Hz intervals in the range [7.5-12.5]. At each sample point, the instantaneous center frequency was determined by computing a weighted average of the attenuation of the signal by each filter. This method resulted in a vector of the same length as the input signal containing the point-by-point estimated center frequency. The variability across this frequency estimate vector was then computed to produce a measure of AFV.

*Automated detection of sleep spindles:* We used the YASA library to do automated spindle detection on all 3 recorded EEG channels entire 30-minute period at the beginning of each session[4]. Spindles were counted in the analyses if they appeared on any of the channels. Spindle statistics including total number, duration, and absolute power were averaged within each condition (Sham or Stim) for each subject, and comparisons were made across conditions. The search frequency range for spindles was set to [12,15]; the minimum spindle duration was set to 0.5 sec and the maximum duration was set to 2 sec, with a minimum distance between successive spindles of 200 ms. Other detection threshold settings were set to default parameters: thresh={'corr': 0.65, 'rel_pow': 0.2, 'rms': 1.5}.

Supplementary Data

*Effect of phase-locked acoustic stimulation on sleep spindles*. In our study, the time from lights-out to the first observable sleep spindle was used as the primary outcome measure. To determine if acoustic stimulation had any observable impact on the occurrence of spindles or spindle characteristics, We performed automated spindle detection on all 3 recorded EEG channels in each session. Using this automated method, the time-to-first-spindle metric was similar between what was previously measured using manual scoring. Manual scorers estimated the weekly average sleep onset times in the Sham condition to be 35.8 ± 17.0 minutes compared to 37.5 ± 22.6 minutes for the automated analysis. For the Stim condition, manual scoring estimated 25.3 ± 15.6 minutes whereas the automated analysis estimated 26.3 ± 17.9 minutes, with a P-value of 0.036 (Supplementary Figure 2a). However, no differences between conditions were found for average spindle duration (Sham = 0.69 ± 0.08 sec; Stim = 0.71 ± 0.06 sec; P = 0.25, Supplementary Figure 2b) or average spindle power (Sham = 1.81 ± 0.53; Stim = 1.94 ± 0.23; P = 0.198, Supplementary Figure 2c).

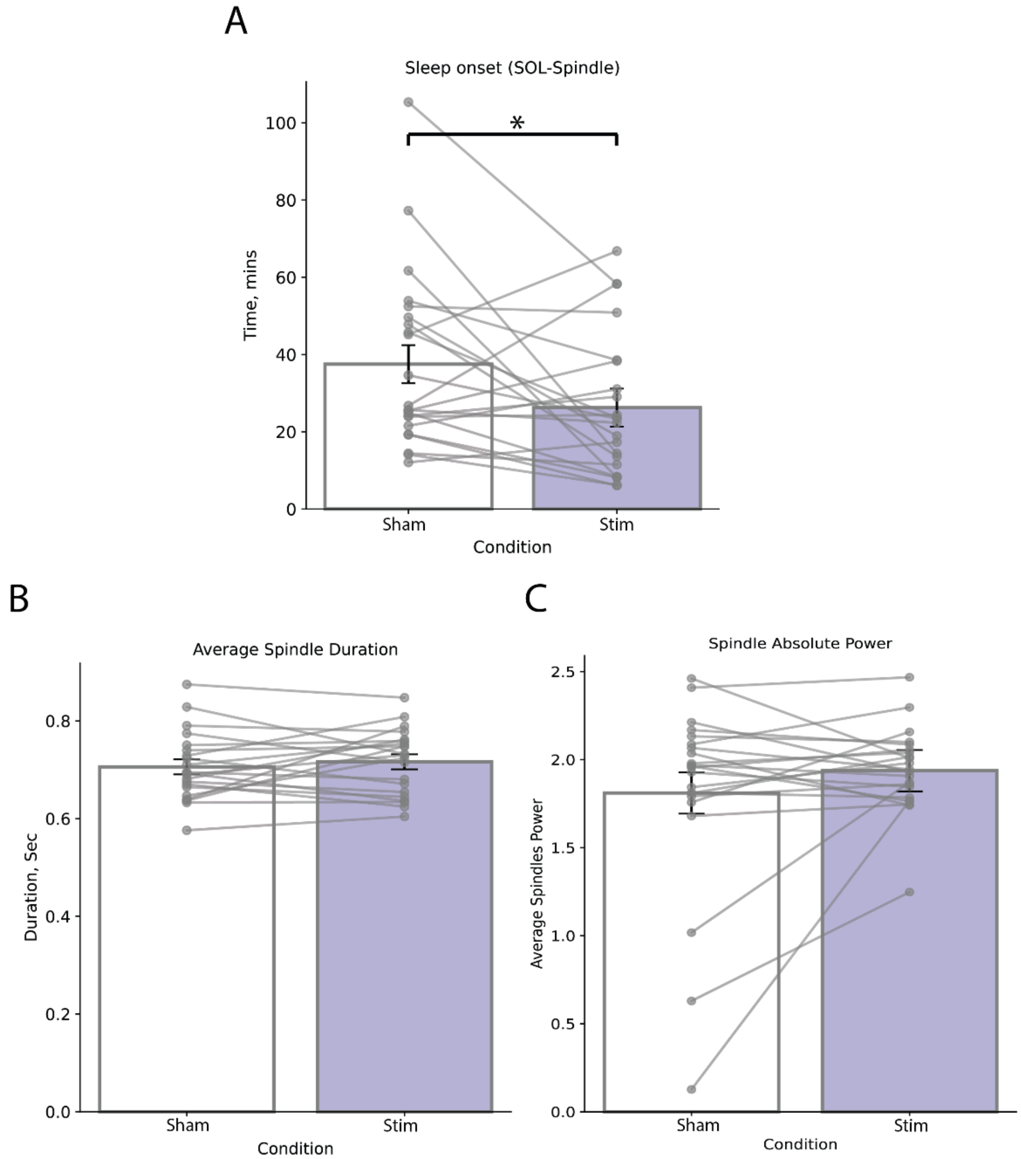

Supplementary Figure 2: Analysis of sleep spindles in Sham compared to Stimulation weeks for each participant. A) Time elapsed between lights-out and the first detected sleep spindle using the automated algorithm. Asterisk indicates P<0.05. Points shown represent the mean values for each subject, and error bars depict the standard error of the mean. B) Average duration of spindles for each subject across conditions. C) Average spectral power of spindles, expressed as median absolute power (in log10 µV^2), calculated from the Hilbert-transform of the signal segment containing each spindle, bandpass filtered between 12 and 15 Hz.

**Supplementary Table 1:** Baseline characteristics of participants excluded during run-in period.

| **Variable** | **Mean ± SD** |
|---|---|
| Age | 34.6 ± 9.32 |
| Gender (female) n (%) | 18 (51.4%) |

| | |
|---|---|
| BMI | 24.8 ± 3.2 |
| Reported time to fall asleep on a typical night (minutes) | 28.7 ± 15.2 |
| Number of caffeinated beverages per day | 1.1 ± 1.0 |
| PSQI score | 10.9 ± 2.8 |
| ISI score | 15.4 ± 4.7 |
| Actigraphy estimated SOL (minutes) | 12.7 ± 10.4 |
| Actigraphy estimated TST (hours) | 6.5 ± 1.4 |
| Actigraphy estimated Eff | 84.6 ± 5.8 |
| Actigraphy estimated WASO (minutes) | 48.1 ± 24.9 |
| Actigraphy estimated nWASO | 24.1 ± 8.7 |
| Subjective SOL (minutes) | 28.7 ± 15.2 |
| Subjective TST (hours) | 6.7 ± 1.6 |
| Subjective WASO (minutes) | 18.8 ± 15.7 |
| Subjective Sleep Quality (0-4) | 2.0 ± 0.5 |

**Supplementary Table 2**: Post-Sleep Morning Survey Part I - Karolinska Sleepiness Scale

| **Karolinska Sleepiness Scale (KSS)**<br>SUBJECT NUMBER:<br>DATE/TIME: | | |
|---|---|---|
| | What is your level of alertness versus sleepiness today? | **CHECK ONE** |
| **1** | Extremely alert (most alert I've ever been) | |
| **2** | Very alert | |
| **3** | Alert | |
| **4** | Somewhat alert | |
| **5** | Neither alert nor sleepy | |
| **6** | Some signs of sleepiness | |
| **7** | Sleepy, but no effort to stay awake | |

| | |
|---|---|
| **8** | Sleepy, with some effort to stay awake |
| **9** | Very sleepy (sleepiest I've ever been) |

Post-Sleep Morning Survey Part II

| **SUBJECTIVE EXPERIENCE QUESTIONS**<br>SUBJECT NUMBER:<br>DATE/TIME: | | |
|---|---|---|
| | | **Answers** |
| **1** | I found the experience of going to sleep pleasant. | Likert scale (1-7) |
| **2** | I found the EEG device use comfortable. | Likert scale (1-7) |
| **3** | The EEG device prevented me from falling asleep. | Likert scale (1-7) |
| **4** | The sound was annoying and/or disturbing. | Likert scale (1-7) |
| **5** | The sound was hypnotic and/or distracting in a good way. | Likert scale (1-7) |
| **6** | The sound was pleasant and/or soothing. | Likert scale (1-7) |
| **7** | The sound helped me fall asleep faster than normal. | Likert scale (1-7) |
| **8** | How often would you use this device if available? | Short Scale (1-5) |
| **9** | I heard the sound changing as I relaxed and fell asleep. | Yes/No |

Use following "Likert" scale to answer Questions 1-7:
1 - Strongly Agree
2 - Agree
3 - Somewhat Agree
4 - Neither Agree or Disagree
5 - Somewhat Disagree
6 - Disagree
7 - Strongly Disagree

Use the following "Short" scale to answer Question 9:
1 – Always (every night)
2 – Often
3 – Sometimes
4 – Rarely

5 – Never

Post-Sleep Morning Survey Part III

| **CONSENSUS SLEEP DIARY (SURVEY OF NIGHTLY SLEEP QUALITY)**<br>SUBJECT INFO:<br>DATE/TIME: | | |
|---|---|---|
| | **Questions** | **Answers** |
| **1** | What time did you get into bed last night?. | Enter the time (ex: 10:15 PM) |
| **2** | What time did you try to go to sleep? | Enter the time (ex: 10:30 PM) |
| **3** | How long did it take you to fall asleep? | (example: 55 min) |
| **4** | How many times did you wake from sleep, not counting your final awakening? | (example: 3 times) |
| **5** | In total, how long did these awakenings last? | (example: 1 hour & 10 minutes) |
| **6** | What time was your final awakening (what time did you wake after you completed your sleep)? | (example: 6:35am) |
| **7** | What time did you get out of bed for the day? | (example: 7:20am) |
| **8** | How would you rate the quality of your sleep? | **Choices:** very poor; poor; fair; good; very good |
| **9** | Comments about last night's sleep (if applicable) | (for example: I have a cold) |

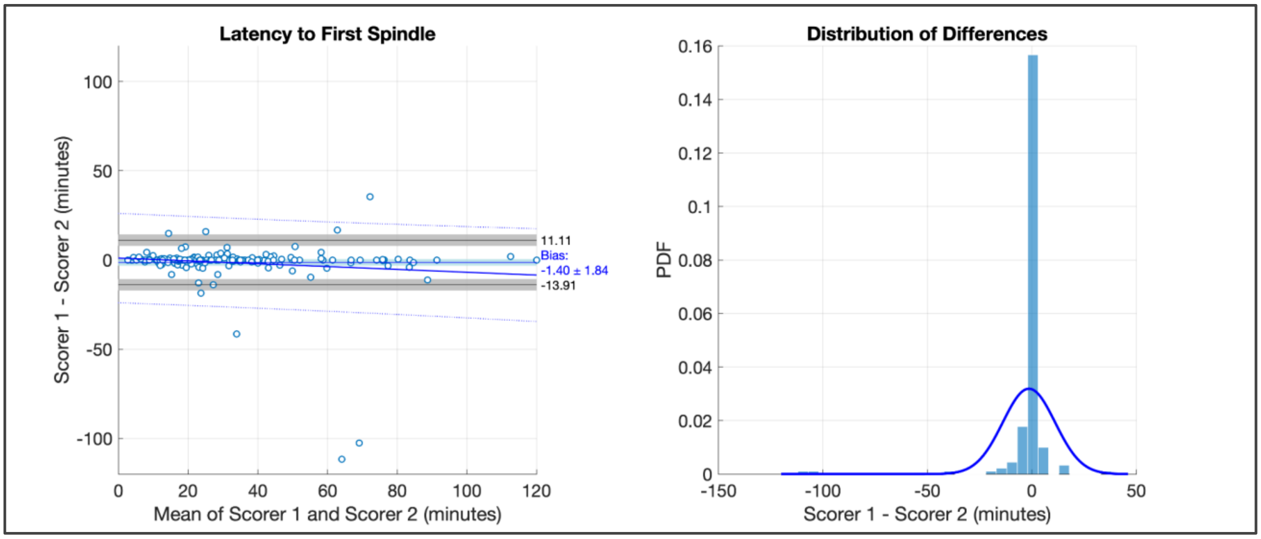

**Supplementary Figure 3:** Bland-Altman analysis comparing Scorers 1 and 2 on estimating the onset latency to the first identifiable spindle.Data logs were flagged if the scored sleep onset time fell outside of the 95% confidence interval of the bias estimate. Flagged data logs were reanalyzed by both scorers working together, and included only if consensus could be reached.

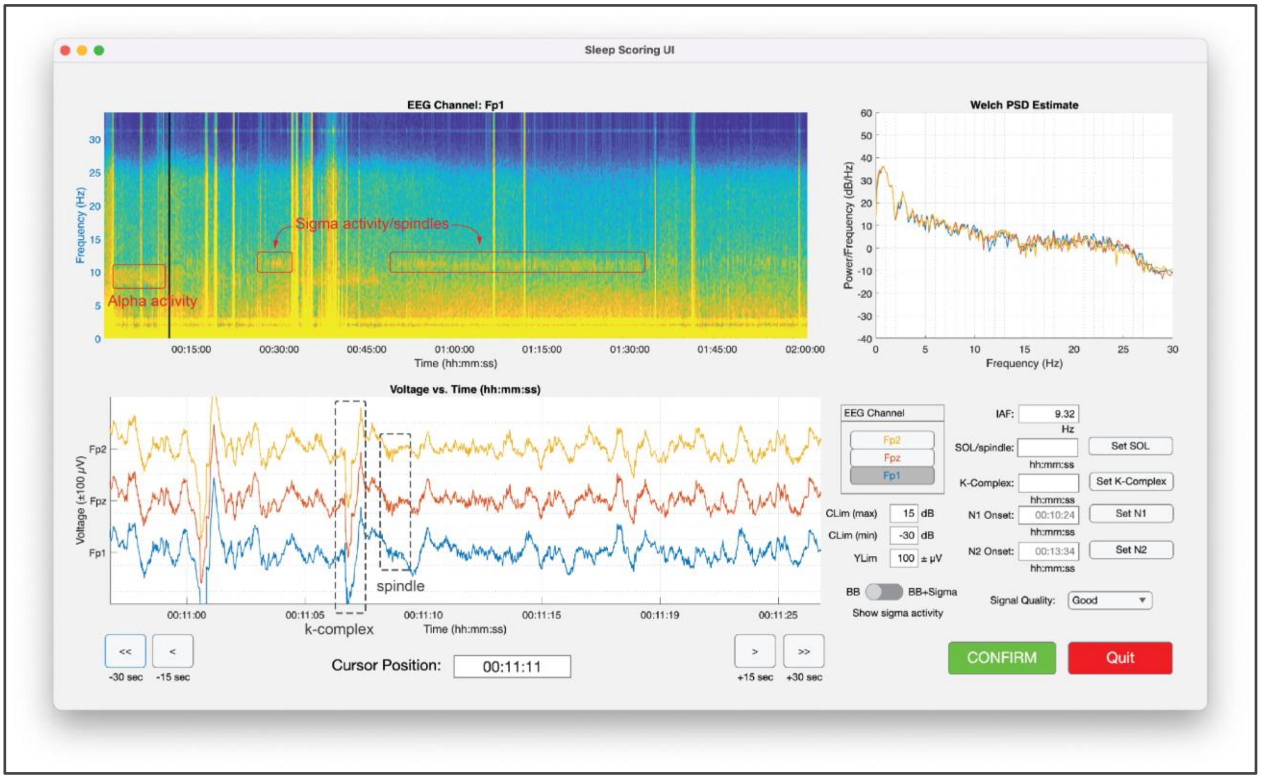

**Supplementary Figure 4:** Sleep Staging User Interface. De-identified EEG data logs were randomly selected for visual scoring. The interface allowed scorers to see the full two-hour spectrogram for any of the three available EEG channels. The solid black vertical cursor on the spectrogram marked the center of a 30-second epoch of the time-voltage representation, which could be advanced in ±15 and ± 30 second intervals.  The power spectrum of the 30-second epoch is shown in the upper right window, where the scorer could manually select the peak of the Individual subject's Alpha Frequency (IAF).  Onset times for key sleep microevents (e.g., time to first spindle, time to first k-complex, onset of N1 and N2 sleep)

were registered with the assistance of the data cursor. Scorers were also instructed to rate the signal quality of the data logs as “Good”,”Fair”,”Poor” or “Unusable”.

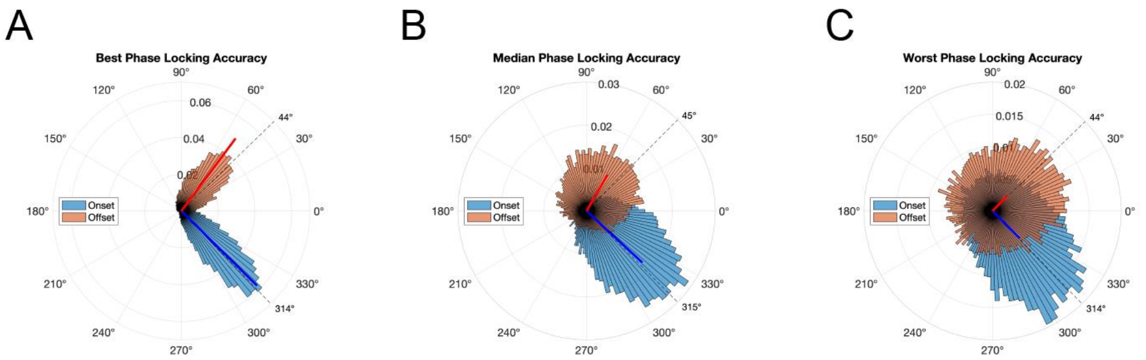

**Supplementary Figure 5**: Phase probability histograms of individual pulse onset and offset events from three data logs representing the best (A), median (B), and worst (C) phase locking accuracy scores. Mean phase angles and phase locking values (PLV) are shown by the solid line vectors. PLVs are proportional to the length of the vector, with perfect phase locking (PLV = 1.00) equivalent to the outermost radius of the polar axis. The dashed lines in each plot represent the targeted alpha phase for pulse onset and offset. Median PLV values for A) were 0.8252 (on), 0.70362 (off); B) 0.59004 (on), 0.32527 (off); and C) 0.29864 (on), 0.16836 (off).

**Supplementary Table 3:** Relationship between Night Order and subjectively reported sleep quality. Values in each box represent the number of nights for the corresponding condition and quality combination. Color saturation indicates the normalized proportion.

**ORDER vs. Sleep Quality**

| ORDER \ SUBJECTIVE SLEEP QUALITY | Very poor | Poor | Fair | Good | Very good |
|---|---|---|---|---|---|
| Night 1 | 0 | 5 | 11 | 16 | 2 |
| Night 2 | 3 | 3 | 12 | 13 | 2 |
| Night 3 | 1 | 3 | 10 | 17 | 1 |
| Night 4 | 0 | 2 | 16 | 12 | 2 |

(Color scale: 0 – 1)

**Supplementary Table 4:** Relationship between Week and subjectively reported sleep quality. Values in each box represent the number of nights for the corresponding condition and quality combination. Color saturation indicates the normalized proportion.

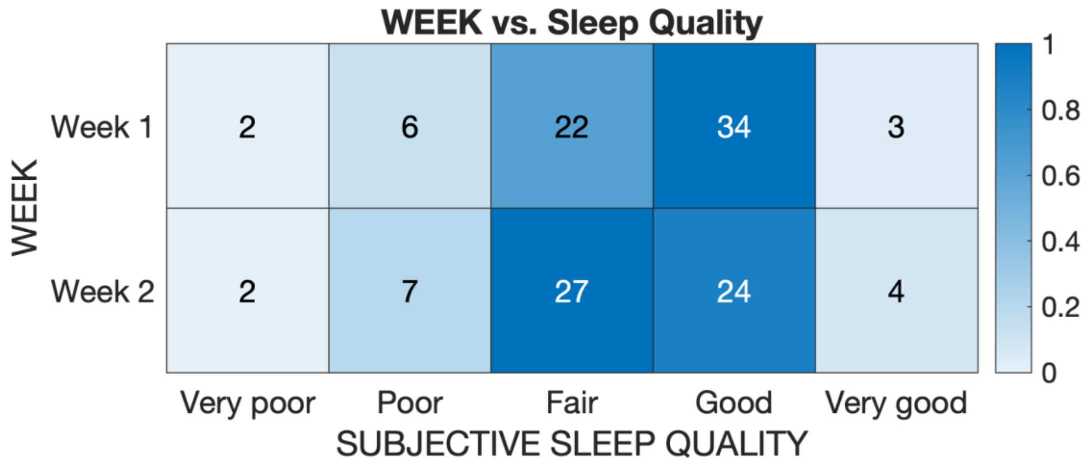

**Supplementary Table 5:** Relationship between Weeknight and subjectively reported sleep quality. Values in each box represent the number of nights for the corresponding condition and quality combination. Color saturation indicates the relative proportion.

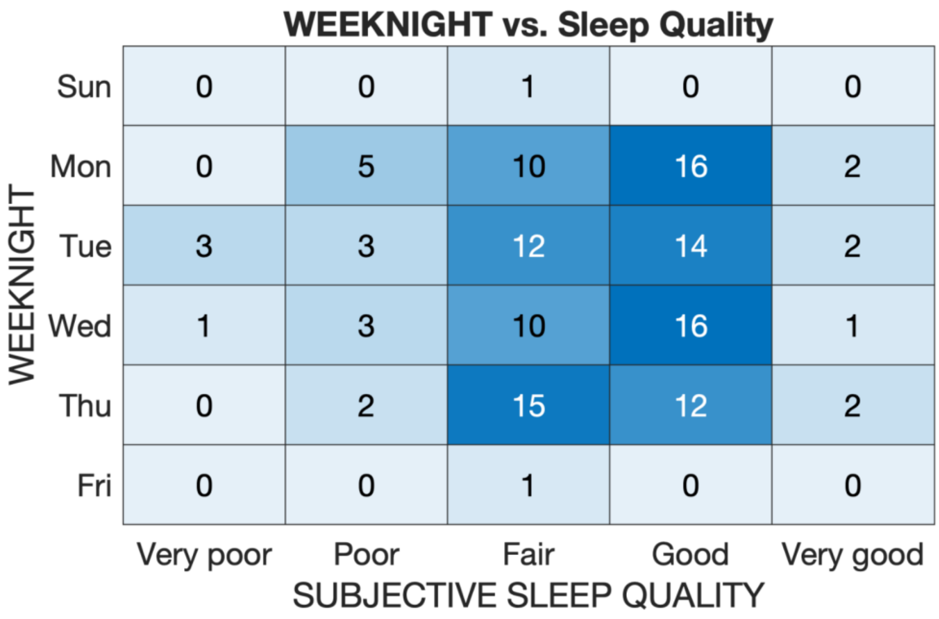
